# SARS-CoV2- infection as a trigger of humoral response against apolipoprotein A-1

**DOI:** 10.1101/2021.02.12.21251298

**Authors:** Sabrina Pagano, Sabine Yerly, Benjamin Meyer, Catherine Juillard, Noémie Suh, Christophe Le Terrier, Jean-Pierre Daguer, Lluc Farrera-Soler, Sofia Barluenga, Giovanni Piumatti, Oliver Hartley, Barbara Lemaitre, Christiane S. Eberhardt, Claire-Anne Siegrist, Isabella Eckerle, Silvia Stringhini, Idris Guessous, Laurent Kaiser, Jerome Pugin, Nicolas Winssinger, Nicolas Vuilleumier

## Abstract

**Aims:** Unravelling autoimmune targets triggered by SARS-CoV-2 infection may provide crucial insights in the physiopathology of the disease and foster the development of potential therapeutic candidate targets and prognostic tools. We aimed at determining i) the association between anti-SARS-CoV-2 and anti-apoA-1 humoral response, ii) their relationship to prognosis, and iii) the degree of linear homology between SARS-CoV-2, apoA-1, and Toll-like receptor-2 (TLR2) epitopes.

**Methods and Results:** Immunoreactivity against different engineered peptides as well as cytokines were assessed by immunoassays, on a case-control (n=101), an intensive care unit (ICU; n=126) with a 28-days follow-up, and a general population cohort (n=663) with available samples in the pre and post-pandemic period. Using bioinformatics modelling a linear sequence homologies between apoA-1, TLR2, and Spike epitopes were identified. Overall, anti-apoA-1IgG levels were higher in COVID-19 patients or anti-SARS-CoV-2 seropositive individuals than in healthy donors or anti-SARS-CoV-2 seronegative individuals (p<0.0001). Significant and similar associations were noted between anti-apoA-1, anti-SARS-CoV-2IgG, cytokines, and lipid profile. In ICU patients, anti-SARS-CoV-2 and anti-apoA-1 seroconversion rates displayed similar 7-days kinetics, reaching 82% for anti-apoA-1 seropositivity. C-statistics (CS) indicated that anti-Spike/TLR2 mimic-peptide IgGs displayed a significant prognostic accuracy for overall mortality at 28 days (CS: 0.64; p=0.02). In the general population, SARS-CoV-2 exposure increased baseline anti-apoA-1 IgG levels.

**Conclusion:** COVID-19 induces a marked humoral response against the major protein of high-density lipoproteins. As a correlate of poorer prognosis in other clinical settings, such autoimmunity signatures may relate to long-term COVID-19 prognosis assessment and warrant further scrutiny in the current COVID-19 pandemic.

## INTRODUCTION

Several lines of evidence point to a SARS-CoV-2-triggered maladaptive immune response as an important determinant of coronavirus disease 2019 (COVID-19) severity (1, 2). Relying on a complex interplay between pathogens and host factors, the B-cell immune-mediated response is characterized by a polyclonal activation leading to the production of numerous antibodies which may cross-react with self-antigens when shared molecular homology between self and non-self-antigens occurs (3-5). Because infection-triggered autoantibodies have been shown to enhance tissue damage and the host inflammatory response, molecular mimicry between self and exogenous epitopes is considered to represent an important mechanism underlying the triad between infectious diseases, autoimmunity, and poorer outcomes (3-5).

Furthermore, concurring emerging data demonstrates that humoral autoimmune mechanisms are frequent in COVID-19, with several autoantibodies being detectable in up to 69% of COVID-19 acute cases (6-9). Sequence/structural homologies between SARS-CoV-2 immunodominant epitopes, the receptor binding domain (RBD) and numerous host antigens have been proposed to underlie such phenomenon (10-12). Recently, we identified three epitopes from the Spike(S) sub-domains S1 and S2, and the C-terminus (c-ter) of Spike as potential immunodominant epitopes of the Spike protein (13), the c-ter of Spike (amino-acid region 1140-1170) having been independently confirmed (14-16). Previous unpublished observations indicated that this Spike c-ter region shares sequence homology with the c-ter of apolipoprotein A-1 (apoA-1), the major protein fraction component of high-density lipoprotein (HDL); while a more proximal RBD region (amino-acid region 455-487), interacting with ACE2 receptor (15), displayed homology with Toll-like Receptor 2 (TLR2). TLR2 engagement and subsequent activation have been shown to be required for autoantibodies against apoA-1 (anti-apoA-1 IgG) to mediate their pro-atherogenic effects (17-20). Because anti-apoA-1 IgGs were shown to represent an independent cardiovascular (CV) risk factor associated with poor prognosis (21-26), to be elevated after certain viral infections (27, 28), and to be preferentially oriented against the c-ter part of apoA-1 (29, 30), we hypothesized that SARS-CoV-2 infection could elicit an anti-apoA-1 IgG response with substantial overlap with anti-SARS-CoV-2 IgG serology.

Therefore, we used bioinformatics modelling coupled with mimetic engineered peptides and we screened three independent COVID-19 adult cohorts for the presence of anti-apoA-1 IgG in, including a case-control (n=101), a prospective intensive care unit (ICU) (n=126), and a general population cohort (n=663). Finally, we performed competition assays to identify the degree of cross-reactivity between anti-apoA-1 IgG and anti-spike IgG.

## RESULTS

### Sequence homology assessment and corresponding Spike mimic peptides synthesis

Capitalizing on prior findings indicating that: i) anti-apoA-1 IgG has to bind to TLR2 due to molecular mimicry in order to generate a pro-inflammatory response by inducing the formation of a TLR2/TLR4/CD14 heterotrimer (18, 19), and ii) that anti-apoA-1 IgG are preferentially oriented against the c-ter of apoA-1 in humans (29, 30); we searched for linear sequence similarities between the Spike protein epitopes (13, 14, 15, 16), apoA-1 and the extracellular part of TLR2.

As shown in Figure 1, these analyses revealed that the amino acid (aa) sequence 1139-1162 of the Spike protein shares sequence homology with the c-ter part of apoA-1 (amino acids 216-243; Figure 1d), and that the aa region spanning 579-587 of Spike protein had a good alignment with those of human TLR2 (aa 456-464; 7 out of 9 amino acids; Figure 1d). Closer inspection of the position of the sequence showed that the secondary structure of the peptide was comparable for both sequence matches (Figure 1d). Specifically, the aa sequence 1139-1162 in Spike is part of an alpha helical bundle with a portion of the peptide sequence unresolved whereas the corresponding apoA-1 sequence is also part of an alpha helical bundle with an unstructured portion. The Spike aa 579-587 peptide is part of a beta turn and continues into a beta pleated sheet. There is remarkable structural homology with the structure of the corresponding peptide on TLR2, namely a beta turn followed by a beta pleated sheet segment (Figure 1d). The same analysis was performed to search for homology between TLR2 or apoA-1 and the N-protein of SARS-CoV-2, using epitopes experimentally detected (15, 16). A good alignment between apoA-1 (aa 131-143) and N (aa 400-412) was identified with 7 identical and 2 similar residues out of 11, however, the lack of structural data for the segment of the N protein precluded further analysis. Likewise, a good match was identified between TLR2 (aa 549-557) and N (aa 217-225) with 5 identical and 2 similar residues in a 8 amino acid stretch, but its structural homology could not be validated. In light of the lack of structural data for these N homology regions, further experimental validation of cross-reactivity was not pursued. Peptide sequences and structures are presented in supplemental figures 1 and 2.

**Fig. 1.**
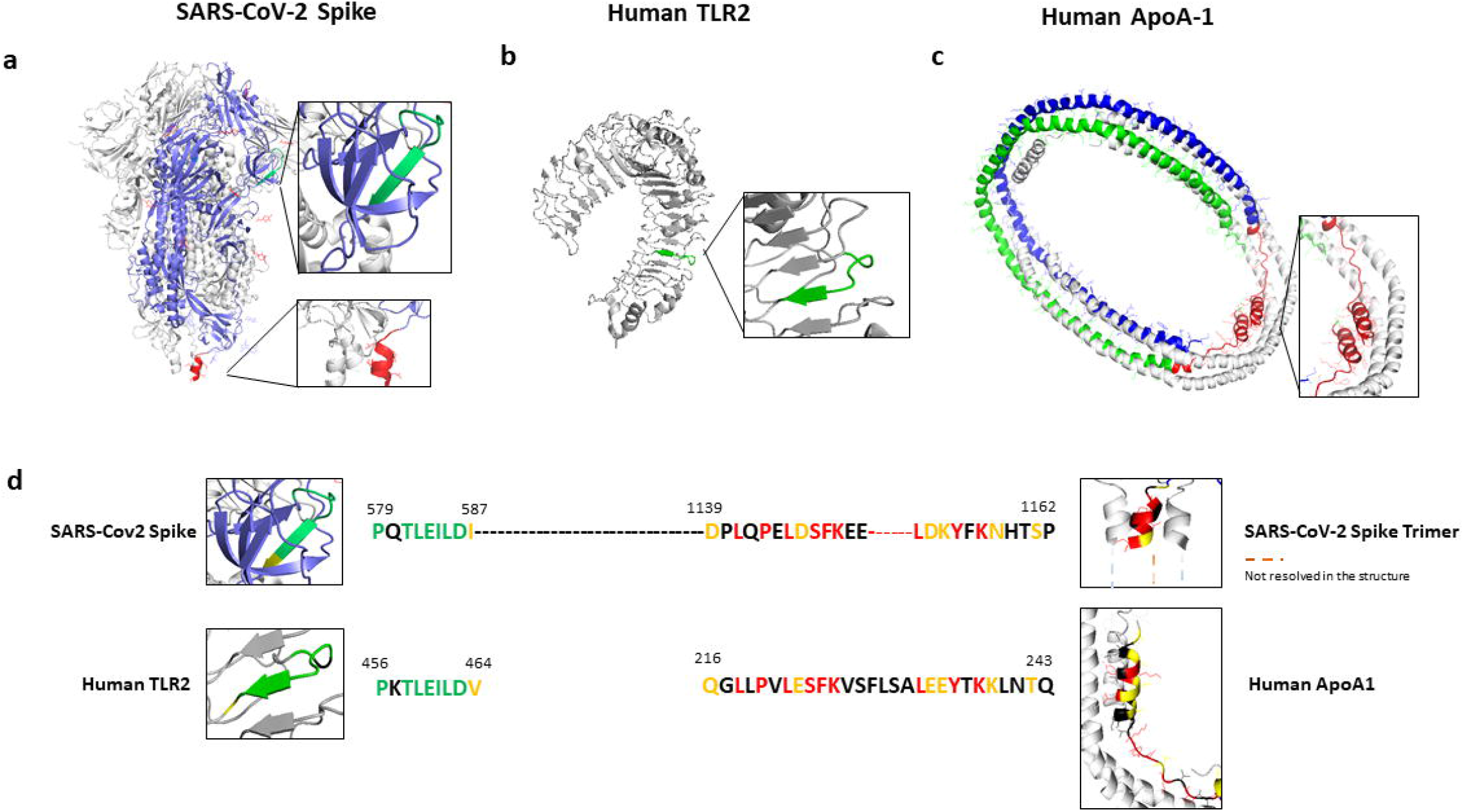
Localization of shared epitopes with apoA-1 and TLR2 on the crystal structure of SARS-CoV-2 Spike protein. Panel a) Crystal structure of SARS-CoV-2 spike protein homotrimer (PDB ID 6VXX). Panel b) Human TLR2 crystal structure (PDB ID 6NIG). Panel c) Human ApoA-1 tetramer crystal structure (PDB ID 1AV1). Epitope sequences conserved between Spike and TLR2 are highlighted in green (a and b) and conserved sequences between Spike and ApoA-1 are represented in red (a and c). Panel d) Sequence alignment of SARS-CoV-2 Spike protein (QIV65088.1) with human TLR2 (H33756AA.1) using Clustal W. Conserved residues are indicated in green and the semiconserved one in yellow. BlastP sequence alignment of SARS-CoV-2 Spike sequence with human apoA-1 (P02647.1). Conserved residues are shown in red and semiconserved (functional equivalent) ones in yellow.

### Anti-SARS-CoV-2 serologies and anti-apolipoprotein A-1 IgG associations

In order to validate our previous bioinformatic findings to humans, we explored the associations between anti-apoA-1 and anti-SARS-CoV-2 serologies on three different cohorts, including a case-control and the ICU cohort, as well as a general population cohort described in figure 2 and 3. In the case-control and ICU cohort, we aimed at replicating the previously reported correlations between anti-apoA-1 IgG with cytokine and lipid profile (20, 31) and explored their possible extension to Spike/apoA-1 and Spike/TLR mimic peptides as an additional orthogonal assessment of closely related serologies. The general population cohort was instrumental to generalize the results observed in acute settings at the population level and to determine whether the pre-pandemic anti-apoA-1 serological status could modulate the anti-SARS-CoV-2 response.

**Fig. 2.**
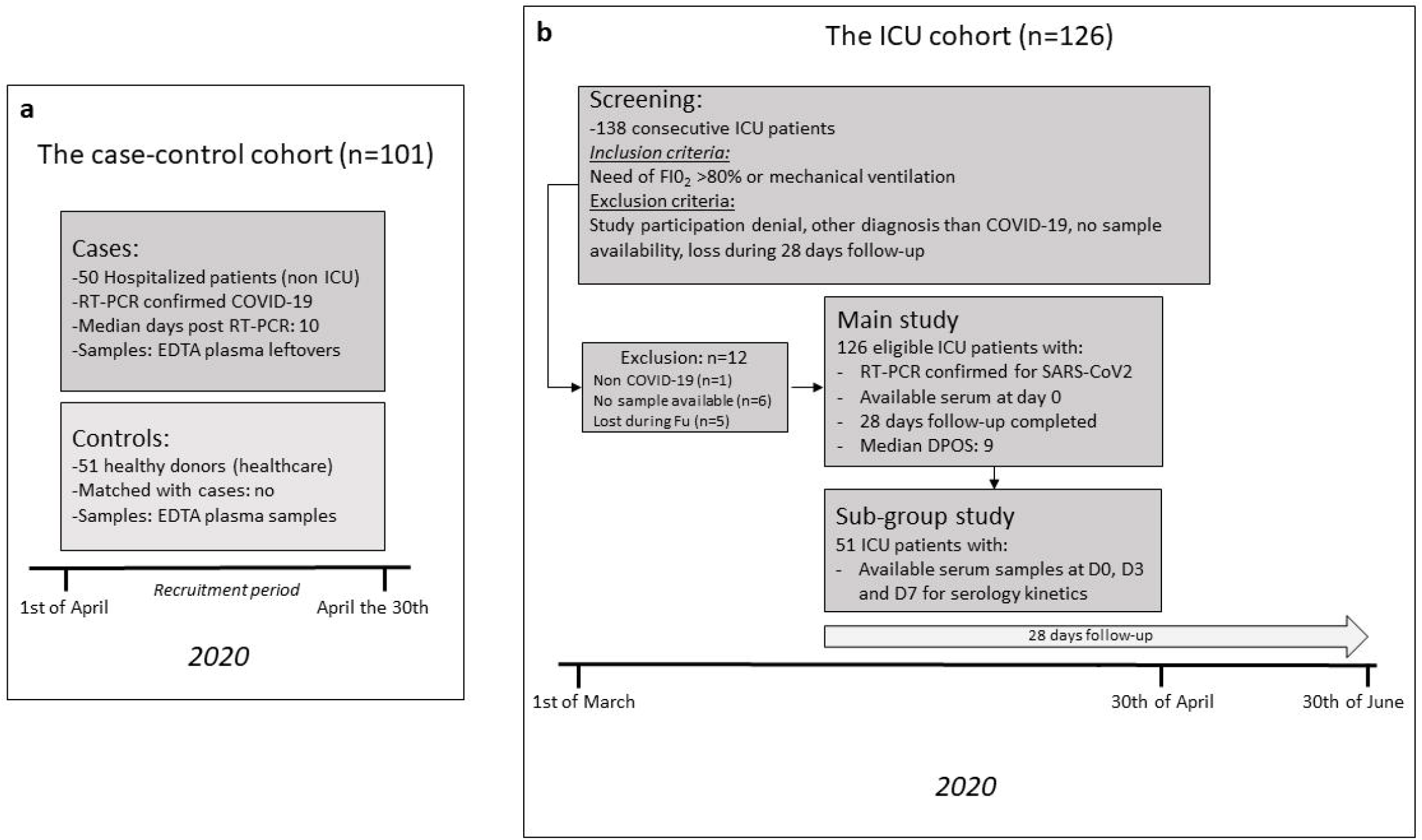
Study flow chart summary of the cohorts included. Abbreviations: DPOS: days post symptom onset ICU: intensive care unit FU: follow-up DPOS: days post symptom onset

**Fig. 3.**
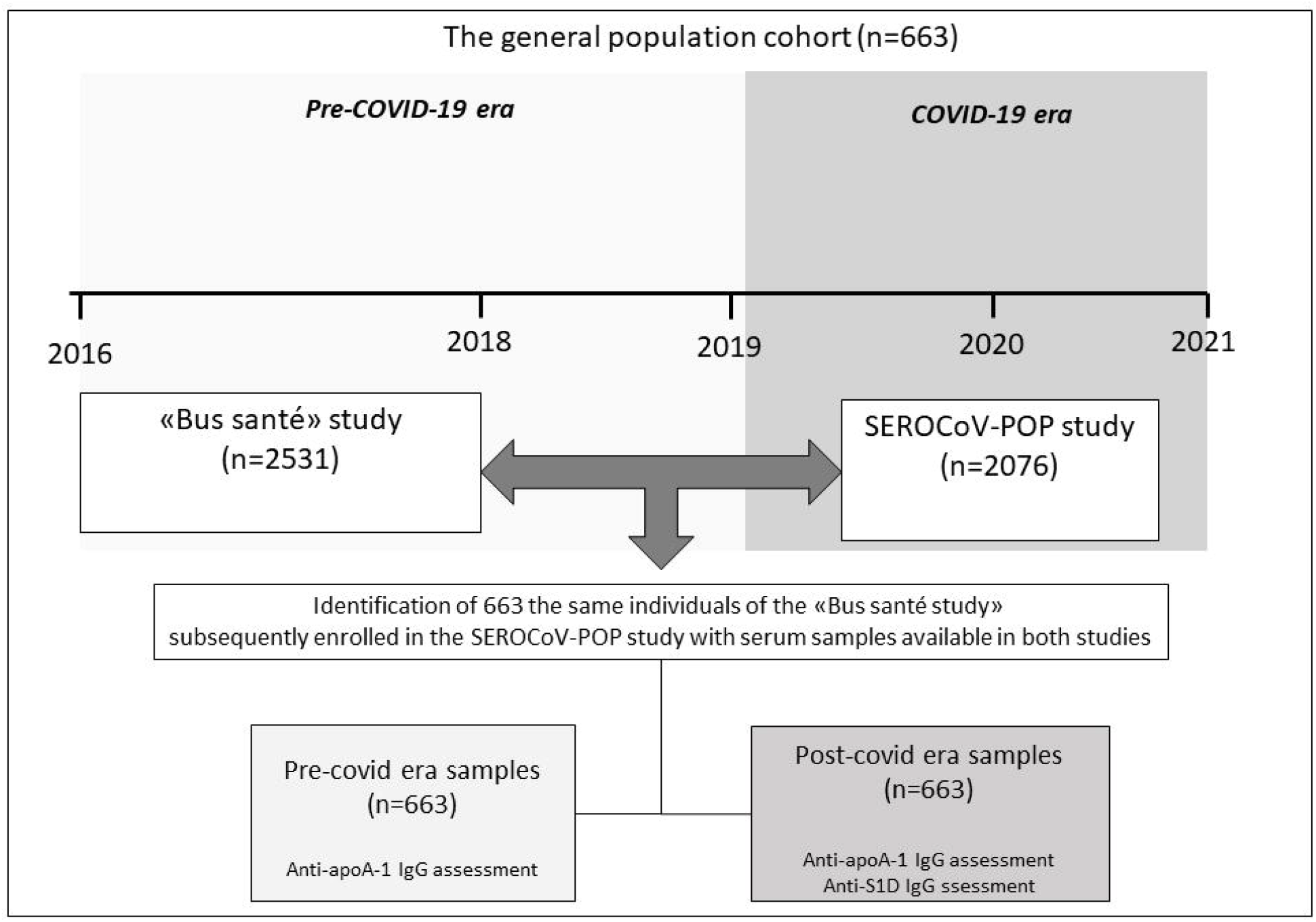
Study flow chart summary of the general population cohort.

### Results from the case-control cohort

The case-control cohort consisted in 50 COVID-19 RT-PCR confirmed cases, and 51 recruitment period-matched healthy donors recruited over the same period (figure 2 panel a). The baseline demographic characteristics of participants are summarized in Table 1. Briefly, RT-PCR confirmed COVID-19 patients were older with an over representation of male genderdisplaying a higher systemic pro-inflammatory state when compared to the healthy blood donors (Table 1). Among COVID-19 patients, the median delay between a positive SARS-CoV-2 RT-PCR diagnostic test and current biomarker assessment was 10 days (IQR 5-15 days). The proportion of patients within each days post-diagnosis subgroup (delta between their molecular testing and serological testing), was 43.5% for 0-6 days (n=20), 30.4% for 7-14 days (n=14) and 26.1% for >14 days (n = 12).

**Table 1.**
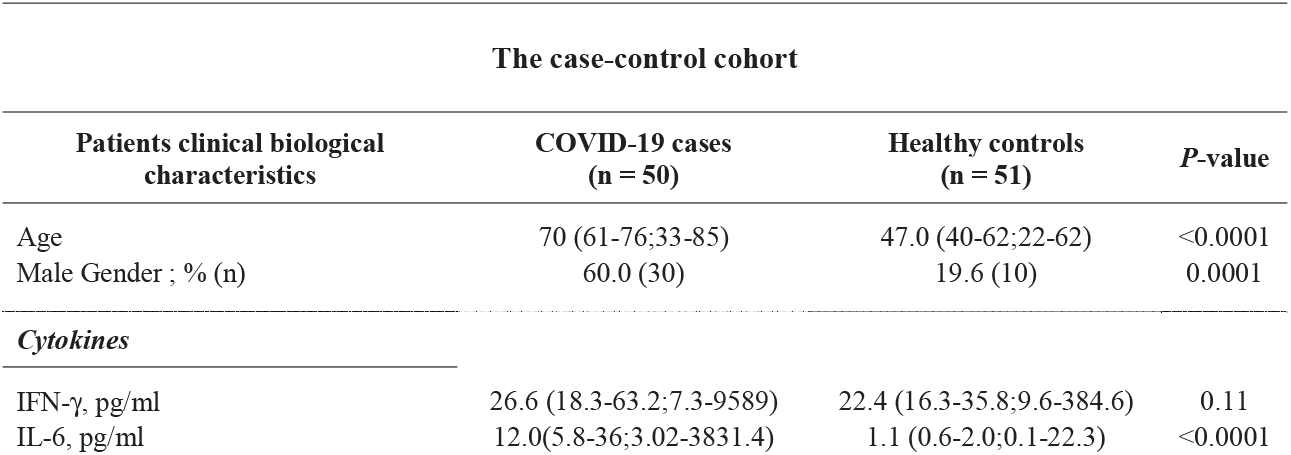
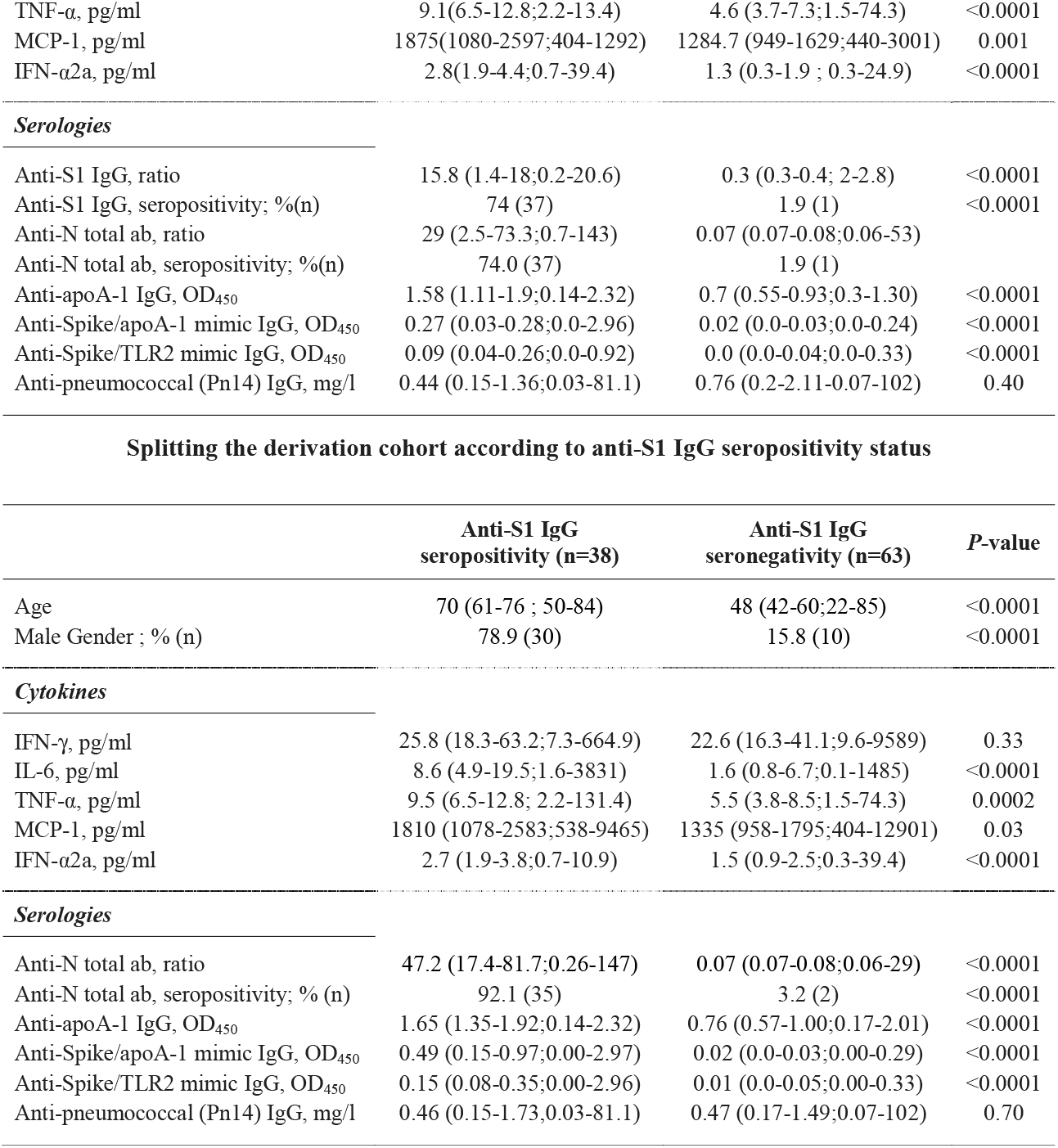
Demographic and biological characteristics of the case-control cohort.

| The case-control cohort |  |  |  |
| --- | --- | --- | --- |
| Patients clinical biological characteristics | COVID-19 cases (n = 50) | Healthy controls (n = 51) | *P*-value |
| Age | 70 (61-76;33-85) | 47.0 (40-62;22-62) | <0.0001 |
| Male Gender ; % (n) | 60.0 (30) | 19.6 (10) | 0.0001 |
| *Cytokines* |  |  |  |
| IFN-γ, pg/ml | 26.6 (18.3-63.2;7.3-9589) | 22.4 (16.3-35.8;9.6-384.6) | 0.11 |
| IL-6, pg/ml | 12.0(5.8-36;3.02-3831.4) | 1.1 (0.6-2.0;0.1-22.3) | <0.0001 |
| TNF-α, pg/ml | 9.1(6.5-12.8;2.2-13.4) | 4.6 (3.7-7.3;1.5-74.3) | <0.0001 |
| MCP-1, pg/ml | 1875(1080-2597;404-1292) | 1284.7 (949-1629;440-3001) | 0.001 |
| IFN-α2a, pg/ml | 2.8(1.9-4.4;0.7-39.4) | 1.3 (0.3-1.9 ; 0.3-24.9) | <0.0001 |
| *Serologies* |  |  |  |
| Anti-S1 IgG, ratio | 15.8 (1.4-18;0.2-20.6) | 0.3 (0.3-0.4; 2-2.8) | <0.0001 |
| Anti-S1 IgG, seropositivity; % (n) | 74 (37) | 1.9 (1) | <0.0001 |
| Anti-N total ab, ratio | 29 (2.5-73.3;0.7-143) | 0.07 (0.07-0.08;0.06-53) |  |
| Anti-N total ab, seropositivity; % (n) | 74.0 (37) | 1.9 (1) |  |
| Anti-apoA-1 IgG, OD<sub>450</sub> | 1.58 (1.11-1.9;0.14-2.32) | 0.7 (0.55-0.93;0.3-1.30) | <0.0001 |
| Anti-Spike/apoA-1 mimic IgG, OD<sub>450</sub> | 0.27 (0.03-0.28;0.0-2.96) | 0.02 (0.0-0.03;0.0-0.24) | <0.0001 |
| Anti-Spike/TLR2 mimic IgG, OD<sub>450</sub> | 0.09 (0.04-0.26;0.0-0.92) | 0.0 (0.0-0.04;0.0-0.33) | <0.0001 |
| Anti-pneumococcal (Pn14) IgG, mg/l | 0.44 (0.15-1.36;0.03-81.1) | 0.76 (0.2-2.11-0.07-102) | 0.40 |

**Splitting the derivation cohort according to anti-S1 IgG seropositivity status**
|  | **Anti-S1 IgG**<br>**seropositivity (n=38)** | **Anti-S1 IgG**<br>**seronegativity (n=63)** | **P-value** |
| --- | --- | --- | --- |
| Age | 70 (61-76 ; 50-84) | 48 (42-60;22-85) | <0.0001 |
| Male Gender ; % (n) | 78.9 (30) | 15.8 (10) | <0.0001 |
| *Cytokines* |  |  |  |
| IFN-γ, pg/ml | 25.8 (18.3-63.2;7.3-664.9) | 22.6 (16.3-41.1;9.6-9589) | 0.33 |
| IL-6, pg/ml | 8.6 (4.9-19.5;1.6-3831) | 1.6 (0.8-6.7;0.1-1485) | <0.0001 |
| TNF-α, pg/ml | 9.5 (6.5-12.8; 2.2-131.4) | 5.5 (3.8-8.5;1.5-74.3) | 0.0002 |
| MCP-1, pg/ml | 1810 (1078-2583;538-9465) | 1335 (958-1795;404-12901) | 0.03 |
| IFN-α2a, pg/ml | 2.7 (1.9-3.8;0.7-10.9) | 1.5 (0.9-2.5;0.3-39.4) | <0.0001 |
| *Serologies* |  |  |  |
| Anti-N total ab, ratio | 47.2 (17.4-81.7;0.26-147) | 0.07 (0.07-0.08;0.06-29) | <0.0001 |
| Anti-N total ab, seropositivity; % (n) | 92.1 (35) | 3.2 (2) | <0.0001 |
| Anti-apoA-1 IgG, OD<sub>450</sub> | 1.65 (1.35-1.92;0.14-2.32) | 0.76 (0.57-1.00;0.17-2.01) | <0.0001 |
| Anti-Spike/apoA-1 mimic IgG, OD<sub>450</sub> | 0.49 (0.15-0.97;0.00-2.97) | 0.02 (0.0-0.03;0.00-0.29) | <0.0001 |
| Anti-Spike/TLR2 mimic IgG, OD<sub>450</sub> | 0.15 (0.08-0.35;0.00-2.96) | 0.01 (0.0-0.05;0.00-0.33) | <0.0001 |
| Anti-pneumococcal (Pn14) IgG, mg/l | 0.46 (0.15-1.73;0.03-81.1) | 0.47 (0.17-1.49;0.07-102) | 0.70 |

As shown in Table 1, COVID-19 patients had higher median levels of all the pro-inflammatory cytokines and serologies tested, with the exception of P14 pneumococcal IgG used as an unrelated serological control. The distribution of serological values between cases and controls is available in Supplementary figure 3. No difference between cases and controls was observed for circulating INF-γ levels. Furthermore, when the cohort was split according to anti-S1 IgG seropositivity status, identical differences were observed (Table 1, bottom panel).

These results were further corroborated by the significant and substantial correlations observed between anti-apoA-1 IgG and both (anti-S1 and anti-N) anti-SARS-CoV-2 serologies, as well as by similar correlations with both anti-Spike/TLR2 and anti-Spike/apoA-1 IgGs (supplemental Table 1). On the other hand, none of the aforementioned antibodies correlated with anti-pneumococcal IgG (Supplementary table 1). Furthermore both anti-SARS-CoV-2 serologies, anti-apoA-1 IgG, anti-Spike/apoA-1 mimic IgG, anti-Spike/TLR2 mimic IgGs displayed similar strength of associations with most of the cytokines measured. Finally, anti-pneumococcal IgG was not correlated to any of the cytokines tested (Supplementary table 1).

### Results from the ICU cohort

To extend and validate these findings in severe COVID-19 disease, we used a cohort of 126 consecutive patients admitted to the ICU for severe COVID-19 disease who completed a follow-up at 28 days, figure 2 panel b. The baseline demographic and biological characteristics of ICU COVID-19 patients are summarized in Table 2. Anti-apoA-1 IgG seropositivity upon ICU admission was found in 26.9% of the ICU patients (34/126), while anti-S1 IgG and anti-N seropositivity was 36.5% (46/126) and 42% (53/126), respectively. When split according to anti-apoA-1 IgG seropositivity status upon patient admission at the ICU, seropositive patients tended to have a more severe *Simplified Acute Physiology Score II* score, a higher number of DPSO at ICU admission, displayed higher median D-dimers levels, anti-S1 IgG levels, anti-Spike/apoA1, and anti-Spike/TLR2 IgG levels, but lower total cholesterol, LDL, and triglycerides levels when compared to anti-apoA-1 seronegative individuals (Table 2). The proportion of anti-S1 and anti-N seroconversions were two–fold higher in anti-apoA-1 IgG seropositive individuals compared to those tested negative for these autoantibodies (55.8% vs 28.2%, *P*=0.006; and 64.7 vs 33.7%; *P*=0.01, respectively). No other significant differences for the remaining parameters were identified between anti-apoA-1 IgG seropositive and seronegative individuals.

**Table 2.** Baseline demographic and biological characteristics of ICU patients and according to anti-apoA-1 IgG serological status.

| **Demographic and biological characteristics** | **Overall (n=126)** | **Anti-apoA-1 IgG seropositive patients (n=34)** | **Anti-apoA-1 IgG seronegative patients (n=92)** | ***P*-value** |
| --- | --- | --- | --- | --- |
| Age, years | 63.5 (57-73; 25-86) | 65.5 (57-73;28-86) | 62.5 (57-72.5; 25-83) | 0.48 |
| Female gender, % (n) | 22.2 (28) | 17.6 (6) | 23.4 (22) | 0.63 |
| BMI, kg/m<sup>2</sup> | 28.1 (25.5-32.1; 15.6-52.4) | 28.3 (25.1-32.8-19.3-52.4) | 28.1 (25.6-31.8; 15.6-50.8) | 0.92 |
| Current smoking, % (n) | 13.5 (17) | 14.7 (5) | 13.0 (12) | 0.78 |
| DPSO and ICU admission | 9.0 (7-11;0-27) | 9.5 (7.5-13.5;3-27) | 9.0 (7.0-10;0-27) | 0.10 |
| ***Comorbidities*** |  |  |  |  |
| Hypertension, % (n) | 47.6 (60) | 44.1 (15) | 48.9 (45) | 0.69 |
| Dyslipidemia, % (n) | 58.3 (35) | 23.5 (8) | 29.3 (27) | 0.66 |
| Diabetes, % (n) | 26.7 (34) | 29.4 (10) | 26.0 (24) | 0.82 |
| Previous IC and or HF, % (n) | 23.8 (30) | 23.5 (8) | 23.9 (22) | 1 |
| Previous stroke, % (n) | 5.5 (7) | 0 (0) | 7.6 (7) | 0.19 |
| Known malignancy, % (n) | 7.9 (10) | 11.7 (4) | 6.5 (6) | 0.46 |
| Chronic kidney disease, % (n) | 7.1 (9) | 5.8 (2) | 7.6 (7) | 1 |
| **Severity upon admission** |  |  |  |  |
| APACHE II score | 22 (14-29; 3-38) | 22 (14-29; 3-38) | 22 (13.5-27.5; 4-37) | 0.61 |
| SOFA score | 6 (4-7;1-11) | 5 (4-7; 2-10) | 6 (4-7;1-11) | 0.44 |
| SAPS II score | 53 (43-65;6-82) | 58 (46-69; 18-78) | 52 (38.5-61.5; 6-82) | 0.09 |
| 28-days mortality, % (n) | 16.7 (21) | 23.5 (8) | 14.1 (13) | 0.28 |
| Length of stay at ICU, days | 16 (10-21;1-48) | 14 (10-18; 1-42) | 16 (10-22; 1-48) | 0.25 |
| Mechanical ventilation, % (n) | 96 (121) | 91.1 (31) | 97.8 (90) | 0.12 |
| **Cytokines and inflammation** |  |  |  |  |
| CRP, mg/l | 154 (92-205;23.1-402.8) | 162.7 (114.1-219.5;23.1-402.8) | 144 (92.4-201;31-311) | 0.35 |
| IFN-γ, pg/ml | 411.1 (112-988.1;152.2-37747) | 308.4 (112.1-691.5;5.2-37747) | 426.7 (175.1-1232.6;175-17778) | 0.22 |
| IL-6, pg/ml | 155.0 (69.1-324.3;6.7-7889.6) | 212.4 (61.4-561.0;6.7-7689.8) | 140.8 (73.3-284.6;13.9-2224.8) | 0.32 |
| TNF-α, pg/ml | 6.7 (4.0-16.6; 0.23-164.5) | 9.3 (4.0-16.9;0.3-71.2) | 6.4 (4.2-14.8;1.6-164.5) | 0.33 |
| MCP-1, pg/ml | 4199 (2509-8488;254-36483) | 3620 (2314-10281;776-36483) | 4300 (2634-7609;254-30256) | 0.92 |
| IFN-α2a, pg/ml | 7.1 (2.3-22.6;0.2-468.5) | 6.3 (1.5-19.1;0.2-92.3) | 8.9 (2.6-30.9; 0.3-468.5) | 0.13 |
| D-dimers; ng/ml | 1531 (961-2476;220-10001) | 1838 (1326-3245;439-10001) | 1433 (863-2174; 220-9999) | 0.03 |
| **Lipid profile** |  |  |  |  |
| Total cholesterol, mmol/l | 2.8 (2.2-3.2;1.1-5.8) | 2.4 (1.9-3.1;1.1-5.2) | 2.9 (2.4-3.3;1.1-5.8) | 0.03 |
| HDL cholesterol, mmol/l | 0.63 (0.47-0.76;0.14-1.95) | 0.52 (0.38-0.61; 0.27-0.88) | 0.66 (0.54-0.79;0.14-1.95) | 0.0001 |
| LDL cholesterol, mmol/l | 1.29 (0.87-1.76;0.00-3.70) | 1.00 (0.62-1.57;0.08-3.70) | 1.43 (1.05-1.79; 0.00-3.58) | 0.02 |
| Triglycerides, mmol/l | 1.64 (1.18-2.24;0.79-4.05) | 1.75 (1.38-2.24;0.81-4.00) | 1.57 (1.15-2.23; 0.79-4.05) | 0.24 |
| **Cardiac biomarkers** |  |  |  |  |
| Hs-cTnT, ng/L | 16.0 (9.7-34.9;3.31-971) | 13.0 (8.1-40.5; 3.7-665) | 17.9 (9.7-33.5;3.3-971) | 0.53 |
| NT-proBNP, pg/ml | 308 (95.6-1015; 15.1-18772) | 363 (110-1437;22-5928) | 278.5 (93.3-909;15.1-18772) | 0.42 |
| **Serologies** |  |  |  |  |
| Anti-S1 IgG, ratio | 0.73 (0.4-1.9;0.3-27.0) | 2.1 (0.5-17.3;0.4-27.0) | 0.6 (0.4-1.15;0.3-21.9) | 0.0009 |
| Anti-S1 IgG, seropositivity; % (n) | 36.2 (46) | 55.8 (19) | 28.2 (26) | 0.006 |
| Anti-N total ab, ratio | 0.46 (0.1-4.70;0.1-36.3) | 2.71 (0.2-11.3; 0.1-36.3) | 0.31 (0.1-1.5; 0.1-29.1) | 0.0004 |
| Anti-N total ab, seropositivity; % (n) | 42.0 (53) | 64.7 (22) | 33.7 (31) | 0.002 |
| Anti-apoA-1 IgG, OD<sub>450</sub> | 0.43 (0.24-0.70;0.00-2.60) | 0.99 (0.83-1.70;0.70-2.60) | 0.33 (0.22-0.44;0.0-0.67) | <0.0001 |
| Anti-Spike/apoA-1 IgG, OD<sub>450</sub> | 0.05 (0.03-0.13;0.00-2.71) | 0.16 (0.06-0.44;0.00-2.71) | 0.04 (0.02-0.07;0.00-1.86) | <0.0001 |
| Anti-Spike/TLR2 IgG, OD<sub>450</sub> | 0.03 (0.02-0.07;0.00-0.77) | 0.06 (0.03-0.10; 0.00-0.24) | 0.02 80.01-005;0.00-0.77) | 0.0003 |
| **Renal function** |  |  |  |  |
| Creatinine; µmol/l | 81.0 (66.5-105; 38-769) | 81.5 (67.5-110.5; 47-173) | 80.5 (65-98, 38-769) | 0.36 |

All continuous variables are expressed as median (interquartile range; and range); \**P* value derived from the comparison between anti-apoA-1 IgG seropositive verse seronegative individuals.
DPSO: days post symptom onset; IC: ischemic cardiopathy; HF: heart failure; APACHE II: Acute Physiology And Chronic Health Evaluation II; SOFA: Sequential Organ Failure Assessment; SAPS: Simplified Acute Physiology Score.

When split according to anti-S1 serological status, anti-S1 IgG seropositive patients were less likely to be known to have chronic kidney disease, they tended to have a shorter length of ICU stay, and they were less likely to require mechanical ventilation (Supplementary table 2). On the other hand, anti-S1 IgG seropositive patients displayed higher number of DPSO before ICU admission, higher medians levels of anti-apoA-1 IgG, anti-Spike/apoA-1, anti-Spike/TLR2 mimic IgG and D-dimers, but lower median levels of INF-γ and INF-α2a. No significant differences were noted regarding the lipid profile and other biological parameters tested (Supplementary table 2). The proportion of anti-apoA-1 IgG seropositivity was increased by 2-fold in anti-S1 seropositive individuals compared to anti-S1 IgG seronegative ones (41.3 vs 18.8%, *P*=0.01; Supplementary table 2). As shown in Supplementary table 3, very similar correlations and strength of associations were found between anti-apoA-1 IgG, anti-S1, anti-N, anti-Spike/apoA-1 and anti-Spike/TLR2 mimic IgG serologies compared to those observed in the case control-cohort. Furthermore, except for INF-γ and INF-α2a, similar observations were made regarding the associations between the different serologies and cytokines levels (Supplementary table3).

**Table 3.** Associations between anti-apoA-1 IgG and anti-S1 IgG status in the general population before and after the first COVID pandemic.

| The general population cohort (identical individuals from the pre and post-COVID era) |  |  |  |  |
| --- | --- | --- | --- | --- |
|  | Pre-COVID era individuals (n=663) | Post-COVID era individuals (n=663) | Post-COVID era individuals split according to S1 serology status (n=663) |  |
|  |  |  | Anti-S1 seropositive individuals (n=50) | Anti-S1 seronegative individuals (n=613) |
| **Anti-apoA-1 IgG** | 25.0 % * | 18.0 % * | 34.0% ** | 16.8% ** |

| *seropositivity in % (n)* | (166) | (120) | (17) | (103) |
| --- | --- | --- | --- | --- |
| *Spearman correlations between anti-apoA-I IgG and anti-S1 IgG levels* | NA | r=0.11;<br>P<0.0001 | r= 0.31; P=0.03 | r=0.04, P=0.30 |

### One-week serological kinetics in ICU patients

In the subgroup of 54 ICU patients for which additional serum samples were available at day 3 and day 7 of ICU admission, significant increases in median values of anti-SARS-CoV-2 and all anti-apoA-1 IgG-related serologies were observed (Figure 4). Accordingly and as expected, anti-SARS-CoV-2 seropositivity rates for both anti-S1 and anti-N serologies were high and reached values above 92% at Day 7. Anti-apoA-1 IgG seropositivity evolution displayed a very similar temporal trend to anti-SARS-CoV-2 serology, reaching 82.4% at Day 7, indicating that anti-apoA-1 IgG serology kinetics closely follow the occurrence of anti-SARS-CoV-2 antibodies over 7 days of severe COVID-19 disease (Supplementary table 4). No such kinetics were observed for anti-P14 pneumococcus IgG.

**Fig. 4.**
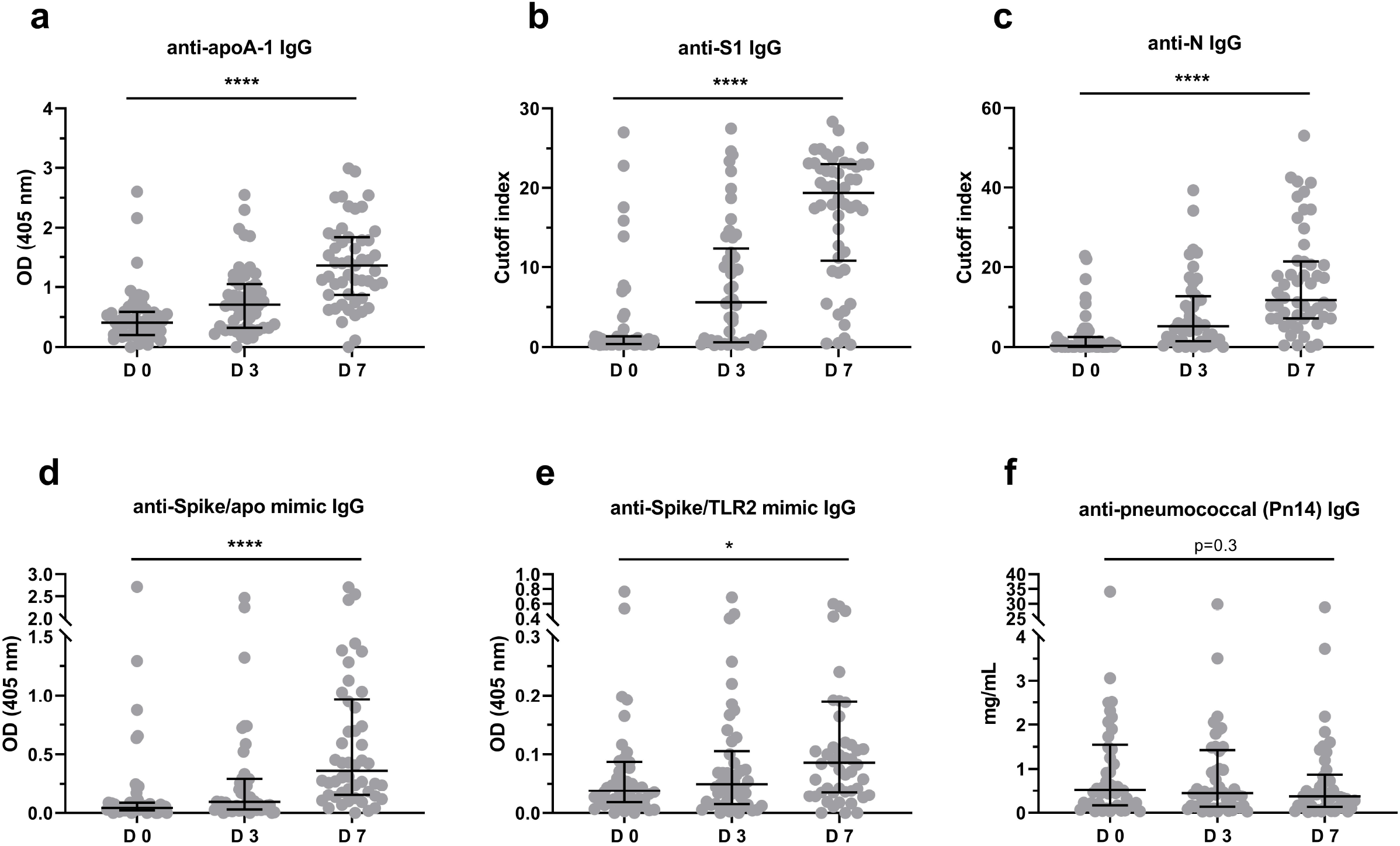
One-week serological kinetics in ICU patients. In **panels a),b),c),d)** and **e)**, ICU patients showed a significant increase (*P*-value of \*\*\*\**P* <0.0001 and *P*=0.025) in antibody titer throughout seven days (days: 0, 3 and 7). In **panel f)** the anti-pneumoccal (Pn14) antibody titer didn’t present any change over time (*P* = 0.3). Results are expressed as median with interquartile range and the Kruskal-Wallis test was used to compare the three groups. Samples were analyzed in duplicate.

### Baseline serologies and ICU patients prognosis

As anti-apoA-1 IgG has been associated with worse overall and cardiovascular prognoses in different studies (21, 22, 25, 26) and because the humoral response against SARS-CoV-2 has been equivocally related to COVID-19 disease severity (32, 33), we investigated their possible prognostic accuracy for overall-mortality at 28 days in ICU patients. ROC curve analyses using continuous biomarker values indicated that the only significant predictor was IgG against the Spike/TLR2 mimic peptide, but only modestly so, with an area under the curve of 0.64. The remaining antibodies were not associated with ICU patients’ prognosis, although a trend was observed for anti-S1 IgG (Supplementary table 5). ROC analyses indicated that the optimal cut-off for anti-spike/TLR2 cut-off would correspond to an arbitrary unit value of 0.11 OD_450_ ratio displaying a sensitivity of 100% (95%CI: 81-100), a negative predictive value of 100% (95%CI: 78-100) a specificity of 17% (95%CI: 13-28) and positive predictive value of 19% (95%CI: 13-28). Given the fact that this cut-off provided no false negatives results, logistic regression analyses could not be performed.

### Results from the general population cohort

Because anti-apoA-1 IgG seropositivity has been shown to concern about one fifth of the general population and to be associated with a poorer prognosis over 5 years (22, 34), our results prompted us to investigate whether such SARS-CoV-2-induced anti-apoA-1 IgG response in acute settings could be replicated in the general population. With this aim, we identified participants recruited in the “Bus Santé” study between 2016 and 2018 and subsequently included in the SEROCoV-POP study (35) during the COVID-19 pandemic. Among these, we identified 663 individuals with available serum samples from both the pre (2016-2018) and post-pandemic (2020) periods (Figure 3 and Supplementary table 6).

The median age of this cohort was 50 years-old (range 24-78), 297 (44.7%) participants were male, while the baseline (pre-pandemic) anti-apoA-1 IgG seropositivity rate was 25.0 % (166/663) and was not associated with any factors commonly ascribed to auto-antibodies such as age, gender or smoking (data not shown). The median time between the first anti-apoA-1 IgG assessment in the pre-COVID-19 period and the anti-S1 IgG plus the second anti-apoA-1 IgG measurement during the post-COVID-19 period (after the first pandemic wave) was 3.2 years (IQR: 2.8-3.6; Range: 2.3-4.3). In the post-COVID-19 period, 7.5% (50/663) were seropositive against SARS-CoV-2 according to anti-S1 IgG levels. The rate of anti-apoA-1 IgG seropositivity, as well as median anti-apoA-1 IgG levels (data not shown) were significantly lower than in the pre-COVID-19 period, despite remaining of the same order of magnitude than previously reported in the general population (21, 22) (Table 3). As shown in Table 3, in the post-COVID-19 samples, a modest but significant correlation was observed between anti-apoA-1 IgG and anti-S1 IgG levels. Furthermore and as shown in Table 3, in the post-COVID-19 period, anti-S1 seropositive individuals displayed higher median anti-apoA-1 IgG levels (0.35 vs 0.57 OD; *P*=0.0002) and higher median anti-apoA-1 seropositivity rates than anti-S1 seronegative individuals (34%.0 vs 16.8%, *P*=0.004). Moreover, the strength of correlation between anti-apoA-1 IgG and anti-S1 IgG in anti-S1 seropositive individuals was of 0.31 (*P* = 0.03), whereas no association between these two serologies was found in anti-S1 seronegative individuals (Table 3). Finally, Cox regression analyses indicated that the pre-COVID-19 anti-apoA-1 IgG status was a significant predictor of post-COVID-19 anti-apoA-1 IgG status (HR: 1.95; 1.52-2.43; *P* <0.0001), irrespective of age, gender and smoking status (HR: 1.57;95%CI: 1.24-1.99; *P*= 0.0001), but did not predict post-COVID-19 anti-S1 seropositivity (HR: 1.30; 95%CI: 0.67-2.54; *P* = 0.44).

### Cross-reactivity and competition experiments

Following the bioinformatics identification of common epitopes between Spike and apoA-1 and the strong association between anti-apoA-1 and anti-SARS-CoV-2 IgG discovered in patients we experimentally try to assess the degree of cross-reactivity between anti-SARS-CoV-2 and anti-apoA-1 IgG with their respective antigens in our ELISA format. As shown in Figure 5a and 5b, we attempted to inhibit the anti-apoA-1 reactivity or the reactivity against the Spike protein incubating an anti-SARS-CoV-2 and anti-apoA-1 IgG double positive sera with increasing concentration of Spike protein and C-ter apoA-1 peptides. As shown in Figure 5a and 5b, neither Spike protein nor c-ter apoA-1 compete for anti-apoA-1 IgG or anti-SARS-CoV-2 IgG signal, respectively. This experimental evidence was furthered by the fact that the polyclonal anti-Spike IgG pre-incubation onto the plate weakly compete for the anti-apoA-1 IgG signal from a pool of anti-apoA-1 IgG/anti-SARS-CoV-2 IgG seropositive individuals (Figure 5c), while anti-apoA-1 IgG pre-incubation did not affect the anti-SARS-CoV-2 IgG signal of the pool of sera patients (Figure 5d). Taken together, these results indicate that, despite the presence of common linear epitopes on Spike, apoA-1, molecular mimicry-driven cross-reactivity between anti-apoA-1 IgG and S1 antigen seemed to be absent.

**Fig. 5.**
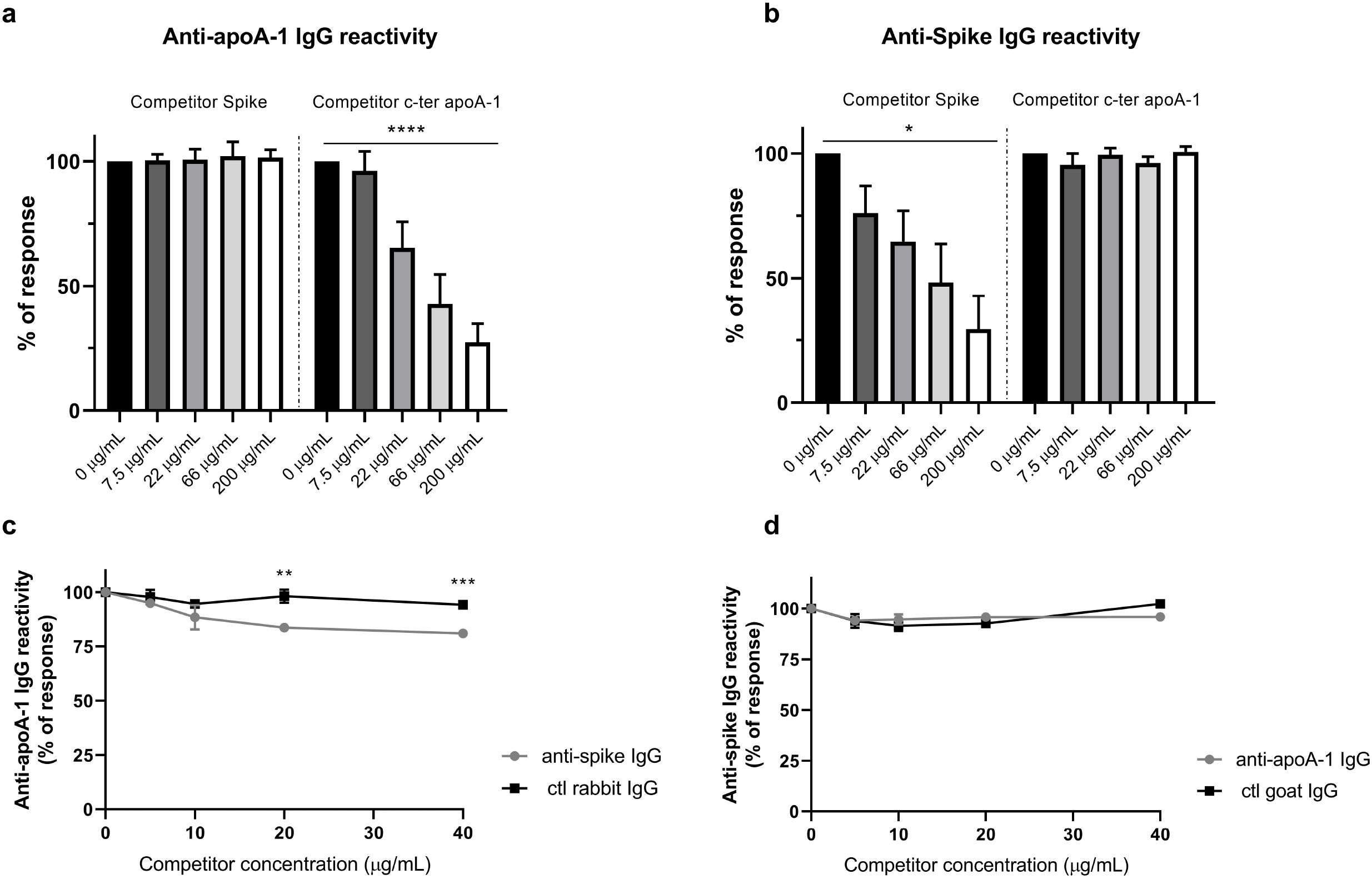
Absence of cross-reactivity between anti-apoA-1 IgG and anti-spike IgG. Panel **a)** and **b)**. Four sera of COVID-19 patients positive for both anti-apoA-1 IgG and anti-spike IgG were pre-incubated with or without spike or c-ter apoA-1 peptides as competitors at the indicated concentrations prior to addition to assay well for anti-apoA-1 IgG or anti-spike IgG measurements. Percentage of maximal ELISA signals were calculated as 100×{[signal in well]−[mean background signal (uncoated well)]}/{[mean maximal signal (no peptide)]−[mean background signal]}. Results are expressed as mean ± SD (n =4). The statistical significance of the signal reduction was calculated by the one-way Anova test : **** *P* < 0.0001 and * *P* = 0.012. Panel **c)**. Polyclonal anti-spike antibodies and not the control rabbit IgGs slightly but significantly compete for apoA-1 binding sites with anti-apoA-1 autoantibodies present in the pooled sera. Results are presented as the mean ± SD of three independent experiments (n = 3). Student’s *T*-test was used to determine the significant difference between the means of the two groups. ** *P* = 0.002 and *** *P* = 0.0002. Panel **d)**. Anti-apoA-1 IgG didn’t compete for Spike protein. Results are presented as the mean ± SD of three independent experiments (n = 3).

To further confirm the absence of such cross-reactivity of IgG against S1 and apoA-1 on anti-S1 IgG results provided in routine diagnosis, 11 samples identified since the beginning of the COVID-19 pandemic that were considered as anti-S1 IgG false-positives, based upon negative recombinant immunofluorescence and negative anti-N serology (35, 36) were tested for anti-apoA-1 IgG. As shown in Supplementary Table 7, none of these eleven samples were found to be positive for anti-apoA-1 IgG.

Taken together, these results do not indicate that anti-apoA-1 IgG could generate analytical interference with anti-S1 serological assay.

## DISCUSSION

The major findings of this study can be summarized by the fact that SARS-CoV-2 infection triggers humoral responses against native and unmodified apoA-1, the major HDL lipoprotein, in up to 80% of severe COVID-19 patients and in up to one third of the infected population. Our study indicates that such phenomenon can be at least partly ascribed to molecular mimicry between linear epitopes of SARS-CoV-2 and those of apoA-1 and TLR2. The c-ter sequence identified in Spike, aa 1139-1162 (15), is known to be conserved in HCoV-OC43 but not amongst other coronaviruses, and was found to share linear homology with aa 216-243 of apoA-1. This c-ter region of apoA-1 structurally corresponds to an alpha helix bundle playing a key role in the cellular cholesterol efflux regulation by ATP-binding cassette transporter A1 (ABCA1) and in HDL maturation (37), and was shown to be preferentially targeted by the polyclonal anti-apoA-1 IgG response (29, 30). Such regional targeting could explain the inverse relationships between anti-apoA-1 IgG and HDL cholesterol reported previously (21, 31, 38), and lately implicated in the disruptive effects of anti-apoA-1 IgG on cellular cholesterol homeostasis (31). The second linear sequence homology identified concerned the aa 579-587 of Spike, located in the S1 domain and close to the receptor binding domain, which has very homology to the aa 456-464 sequence of TLR2. This sequence is part of the leucine reach repeats (LRR) ectodomain of TLR2, known to be key for proper pathogen-associated or damage-associated molecular pattern recognition and TLR2’s function (39). The engagement of this region by anti-apoA-1 IgGs due to sequence homology with apoA-1 (20) is believed to mediate their pro-atherogenic response (18-20, 31), affecting myocardial necrosis and mice survival through specific signaling pathways (18-20, 40). Taken together, these results indicate that these two SARS-CoV-2 epitopes previously identified as potential T cell and B cell epitopes (13, 14, 15, 41), share linear sequence homologies with host antigens playing key roles in inflammation and lipid homeostasis regulation, and that the COVID-19-induced humoral response may elicit pathogenic antibodies potentially cross-reacting with numerous autoantigens due to pathogenic B cell activation (42). While the linear epitope homology between apoA-1, TLR2 and the N antigen could not be validated because of a lack of structural information, significant and substantial associations were still observed between the anti-N and anti-apoA-1-related serologies. These results could also be explained by the possible existence of common conformational epitope(s) that our bio-informatics and epitope mapping systems could not detect, and/or by the likely existence of additional mechanisms, such as intermolecular epitope spreading, allowing the initial targeted humoral response to quickly broaden to antigens other than the inducing epitope (43).

They are three main potential clinical implications of the present findings. Firstly because, anti-SARS-CoV-2 IgG responses against the regions containing these two epitopes were previously shown to correlate with COVID-19 severity (15) and because the presence of anti-apoA-1 IgG has been predictive of poorer outcomes (21, 22, 25, 26, 34, 38, 40), these findings may have possible implications for COVID-19 patients’ prognosis and risk stratification. Indeed, corroborating these previous observations, our results indicate that SARS-CoV-2-induced IgGs against aa 456-464 of TLR2 (and cross-reacting with anti-apoA-1 IgG (20)) could significantly predict 28-day mortality in ICU patients despite a limited sample size, with a NPV reaching 100% using a post-hoc defined cut-off. Although requiring further prospective validations, these preliminary results indicate that assessing such autoantibodies on top of existing stratification tools could pave the way for enhanced risk stratification and resource allocation optimization, a major focus point in severe COVID-19 disease. Secondly, from an analytical standpoint, our results indicate that despite the intimate relationship between anti-apoA-1 and anti-S1 serology and the existence of linear sequence homology between defined apoA-1 and S1 epitopes, our results indicate that the risk of potential analytical interference between these serologies can safely be ruled-out. Thirdly, if the molecular mimicry hypothesis underlies the induction of anti-apoA-1 IgG after a natural SARS-COV-2 antigens exposure, knowing whether such phenomenon could be induced by other antigen exposure needs to be addressed.

From a physiopathological point of view, these results provide innovative perspectives. First of all, these results extend the coverage of virus-mediated anti-apoA-1 IgG induction to SARS-CoV-2, as previously shown for HCV and HIV (27, 28). The inverse associations reported here between anti-apoA-1 IgGs and HDL levels are similar to what has been observed in HCV (27), and are reminiscent of the concept that HCV could hijack the scavenger receptor B-1 (SR-B1)-mediated HDL uptake to infect hepatocytes (27, 44) which has recently been transposed to SARS-CoV-2 by the recent demonstration of Wei and colleagues who identified SR-B1 as an additional receptor facilitating the SARS-CoV-2 entry into cells (45). SR-B1 being the canonical apoA-1/HDL receptor involved in reverse cholesterol efflux and HDL maturation, our results suggest that the molecular mimicry-based anti-apoA-1 IgG response in COVID-19 may concur with other established inflammatory factors to explain the low HDL and apoA-1 levels reported previously in COVID-19 (46, 47). Secondly and along the same line, our results lend further weight to the fact that host lipid metabolism may play an important role in COVID-19 severity by modulating the intensity of the immune response. Lee W et al. (48) recently demonstrated that COVID-19 activates regulatory element binding protein-2 (SREBP-2), a key transcription factory lying at the cross-roads of inflammation modulation and cholesterol biosynthesis. Given the requirement of cholesterol biosynthesis for SARS-CoV-2 budding-driven exocytosis, any factor modulating SREBP-2 pathway activation may influence the course of COVID-19 disease (48). In this respect, recent findings in human macrophages indicate that anti-apoA-1 IgGs increase the expression of SREBP-2 in a TLR2/4-dependent manner, culminating into enhanced foam cell formation, the hallmark of atherogenesis (31), through anti-apoA-1 IgG-dependent ACAT activation leading to the redirection of cellular cholesterol towards intracellular esterified cholesterol pools and decreased membrane free-cholesterol content (31). Because membrane free-cholesterol is a key regulator of membrane ACE2R trafficking into dedicated lipid rafts for optimal SARS-CoV-1 endocytosis (49), such results suggest that by modulating the membrane free-cholesterol content anti-apoA-1 IgG may influence the course of COVID-19. Although devoid of any experimental demonstration, such a hypothesis remains to be further investigated.

We acknowledge several limitations of the present work. Firstly, we limited our analyses to autoantibodies directed against apoA-1 and did not consider other autoantibodies of possible relevance in COVID-19, such as anti-phospholipid or anti-platelet 4 auto-antibodies (50, 51). Secondly, in the context of the recent SARS-COV-2 variants unknown during the first epidemic wave, we could not asses the possible impact of such variant on the anti-apoA-1 IgG response. However, as the linear sequence homologies identified between apoA-1 and Spike did not contain the characteristic epitope regions of the three main variants of concern (VOC) in Europe (United Kingdom, Brazilian and South-African strains), it is unlikely that our results could be specific of a defined and currently existing SARS-CoV-2 strain. Thirdly, the relatively limited number of patients enrolled in this study impeded our ability to complete the survival analyses with the desired level of details regarding the prognostic value of anti-Spike/TLR2, which needs replication in larger prospective cohorts. Finally, our analysis used linear epitope mapping and doesn’t consider potential conformational epitopes which may further contribute to the association between the anti-apoA-1 and anti-N serology. Nevertheless, as those two linear epitopes are exposed to the immune system regardless of the conformation of Spike (monomeric or trimeric) (13, 14, 15), potential conformational issues are unlikely to have blunted the present conclusions. Lastly, if our results indicate that an acute exposure to SARS-CoV-2 rapidly increases the anti-apoA-1 IgG response, they do not allow inferring any conclusions about the possible longer term persistence of anti-apoA-1 IgG levels after COVID-19 disease.

In conclusion, this report shows for the first time that in a substantial proportion of SARS-CoV-2 exposed individuals, a marked humoral autoimmune response against the major lipoprotein of HDL occurs partly due to molecular mimicry between Spike and apoA-1 epitopes. Knowing whether the pre or co-existence of anti-apoA-1 IgG may modulate the course of COVID-19 disease remains uncertain. However, as correlates of poorer prognosis in different settings, a better understanding of the possible clinical relevance of COVID-19-induced autoimmune biological signatures is warranted in the current COVID-19 pandemic and ongoing vaccination programs.

## METHODS

### Sequence homology analyses, Spike-apoA-1 and Spike-TLR2 mimic peptides synthesis

Homologies between Spike and Nucleocapsid epitopes (13-16), TLR2 or c-ter apoA-1 were assessed using Clustal Omega and BlastP sequence alignment. Identified sequences were then mapped onto the structure of the respective proteins using PDB ID 6VXX for SARS-CoV-2 Spike, 6WJI for SARS-CoV N, 6NIG for human TLR2 and 1AV1 for human ApoA-1 to evaluate the surface accessibility of the peptide sequences as well as secondary structure similarities. Sequence homologies were identified between amino acids (aa) 579-587 of Spike and aa 456-464 of TLR2 as well as aa 1139-1162 of Spike and aa 216-243 of apoA-1, but no probable structural homologies between N antigen, apoA-1 and TLR2 were identified.

After sequence homology assessment, the peptides corresponding to the epitopes of Spike and the corresponding peptide sequences from TLR2 and apoA-1 were chemically synthesized by automated synthesis on an Intavis peptide synthesizer as previously described (13). The Spike/TLR2 mimic peptide comprised 13 aa (aa577-588 from SARS-CoV-2 Spike protein + a N-terminus glycine) with the following sequence: GRDPQTLEILDIT, and a molecular weight (MW) of 1469 Daltons. The Spike-apoA-1 mimic peptide consisted of 24 aa (aa1139-1162 from SARS-CoV-2 Spike protein + a N-terminus glycine), with the following sequence: GDPLQPELDSFKEELDKYFKNHTSP, and a MW of 2933 Daltons. The peptides were purified by HPLC and the respective masses were verified by mass spectroscopy (MALDI), and the peptides were reconstituted in water at a concentration of 2mg/mL. The purity of both peptides was 90-95% according to HPLC and MALDI-MS analysis.

### Study populations and sample collection

#### The case-control cohort

Similar to our previous publication (36), the case-control cohort consisted of 50 consecutive real-time RT-PCR confirmed COVID-19 cases hospitalized at the University Hospitals of Geneva (HUG), and 51 recruitment period-matched blood samples from asymptomatic blood donors, enrolled between the 1^st^ to the 30^th^ of April 2020 (first epidemic wave in Geneva). Anti-apoA-1 IgGs levels being independent of age and gender (21), matching for these factors was not considered. Our institution being transformed in a COVID hospital, considering patients hospitalized during the same period for other reason than COVID-19 disease as a control group was not feasible. In accordance with our institution ethical committee and national regulations, irreversibly anonymized leftovers of EDTA plasma were used, in accordance with the Declaration of Helsinki. The study objectives of this case-control cohort were: i) to evaluate the associations between anti-Spike SARS-CoV-2, anti-apoA-1 serologies, as well as with anti-mimetic peptides IgGs; and ii) study their respective associations with systemic cytokines levels.

#### The ICU cohort

The ICU cohort consisted of 126 consecutive patients admitted to the intensive care unit of HUG, between the 1^st^ of March and the 30^th^ of April 2020 for which serum sample leftovers were available from within 24 hours of admission to the ICU of patients who also completed a 28-day follow-up. This study was approved by the Geneva Ethic Council for Research (CCER) (Protocole Nr: CCER 2020-00917, Geneva, Switzerland). The written informed consent was obtained by patients prior enrollment in accordance with the Declaration of Helsinki. Informed consent was obtained either from the patient or from the next-of-kin, or it was deemed to have been given by default, unless a formal opposition was formulated.

The inclusion criteria consisted of admission to the ICU with acute respiratory failure due to SARS-CoV-2 infection requiring mechanical ventilation or a fraction of inspired oxygen (FiO_2)_ >80% between 1^st^ of March and the 30^th^ of April 2020. Exclusion criteria consisted of documented refusal to participate to the study, a negative SARS-CoV-2 RT-PCR, serum sample unavailability upon ICU admission, or incomplete 28-day follow-up. All the relevant medical information, including days post symptoms onset (DPSO) and ICU admission, as well as the 28-day follow-up, were retrieved in the electronic patient medical files by ICU ward physicians, which were blinded to the biochemical analyses. During the inclusion period, 138 patients were admitted to the ICU. Among them, one patient was admitted for a non-COVID-19 pneumonia, 5 patients did not complete the 28-day follow-up due repatriation to their country of origin, and six additional patients did not have available serum leftovers, leaving 126 ICU COVID-19 patients available for the analysis.

The primary biological objective of the study was to explore the associations between anti-SARS-CoV-2 and anti-apoA-1 IgG responses, as well as between the IgG responses against two mimic peptides, including Spike-apoA-1 mimic, and Spike-TLR2 mimic. The secondary biology objectives consisted of exploring the associations between the aforementioned serologies with inflammation biomarkers and the lipid profiles. The third biological objective was to determine the short-term serological kinetics in a subgroup of 51 patients for which additional samples at day 3 and day 7 of ICU admission were available. The only primary clinical study endpoint was overall mortality at 28 days.

#### The general population cohort

This cohort consisted of individuals initially enrolled in the BUS-Santé study in the pre-COVID-19 era (n=2531; recruited in years 2016-2018) which were then subsequently enrolled in the SEROCoV-POP study (n=2076), which aimed at assessing the seroprevalence of anti-SARS-CoV-2 antibodies in the Geneva cantonal population during the first pandemic wave (35). Briefly, for these 2076 individuals, pre-pandemic serum samples stored at -80°C after being collected between February 2016 and January 2018 were available from 663 individuals to assess the anti-apoA-1 IgG response. This study was approved by the Geneva Ethic Council for Research (Protocole Nr: CCER16-363), and written informed consent was obtained by patients prior enrollment in accordance with the Declaration of Helsinki. Further details of the initial SEROCoV-POP have been published previously (35). The two predefined study objectives were to assess at the population level: i) the impact of SARS-CoV-2 infection on anti-apoA-1 IgG levels, and ii) the impact of pre-pandemic anti-apoA-1 serological status on the subsequent anti-SARS-CoV-2 humoral response.

The study flow chart summary of the three cohort is presented in Figures 2 and 3.

#### Sample processing

Whole blood samples were obtained: at the time of donation for healthy donors; at the time of hospital and ICU admission, respectively for cases of the derivation and validation cohorts; and at the time of outpatient recruitment for the general population cohort. Plasma and serum were generated from whole blood via centrifugation. Aliquots were frozen at −80 °C until analysis. Nasopharyngeal swabs for cases of the derivation and the validation cohort, were obtained at the time of hospital and ICU admission.

### SARS-CoV-2 RT-PCR analyses

As previously reported (35, 36), SARS-CoV-2 RT-PCR was performed according to the manufacturers’ instructions on various platforms, including initially in house method using the BD SARS-CoV-2 reagent kit for BD Max system (Becton, Dickinson and Co, US) and Cobas 6800 SARS-CoV-2 RT-PCR (Roche, Switzerland).

### Anti-SARS-CoV-2 against Spike 1 domain IgG assessment

As previously reported (35, 36), we used the Euroimmun IgG enzyme-linked immunosorbent assays (ELISA) (Euroimmun AG, Lübeck, Germany # EI 2606-9601 G; CE-marked) to assess SARS-CoV-2 IgG serology against the S1-domain of the Spike protein (anti-S1 IgG). EDTA-plasma (derivation cohort) and serum (validation cohort) were diluted at 1:101 according to the manufacturer’s instructions. Results of patient sample’s immunoreactivity are expressed as the ratio of the optical density at 450 nm (OD_450_) divided by the calibrator’s OD_450_. The ratio is interpreted as follows: OD_450_ ratio: <0.8 = negative; ≥0.8 and < 1.1 = indeterminate; ≥1.1 = positive. For the purpose of this study, indeterminate results were considered as negative (36). Inter-assay variation was 15.6% for IgG at a ratio of 2.09 (n=17).

### Assessment of total antibodies against N antigen of SARS-CoV-2

Total antibodies against the N antigen of SARS-CoV-2 were measured on a Cobas e801 analyzer (Roche Diagnostics, Rotkreuz, Switzerland) according to the manufacturer’s instructions. Results are reported as numeric values in form of a cut-off index (signal sample/cutoff or signal calibrator ratio) and are considered as positive when equal to or above 1. Inter-assay variation was 14.3% at a ratio of 2.97 (n=17).

### Anti-apoA-1 IgG assessment

Anti-apoA-1 IgGs were measured as previously described (21, 22, 25-28, 38), using frozen EDTA plasma (case-control cohort) and serum (ICU and general population cohort). Maxisorp plates (Nunc™, Roskilde, Denmark) were coated with purified, human-derived delipidated and unmodified apoA-1 (20 µg/mL; 50 µL/well) for 1 h at 37 C. After being washed, all wells were blocked for 1 h with 2% bovine serum albumin (BSA) in a phosphate buffer solution (PBS) at 37 °C. Participants’ samples were also added to a non-coated well to assess individual non-specific binding. After six washing cycles, a 50 µL/well of signal antibody (alkaline phosphatase-conjugated anti-human IgG; Sigma-Aldrich, St Louis, MO, USA, ref: A-3150), diluted 1:1000 in a PBS/BSA 2% solution, was added and incubated for 1 h at 37 C. After washing six more times, phosphatase substrate p-nitrophenyl-phosphate-disodium (Sigma-Aldrich, St Louis, MO, USA) dissolved in a diethanolamine buffer (pH 9.8) was added and incubated for 30 min at 37 °C (Molecular Devices™ Filter Max F3, Molecular Devices, San Jose, CA, USA). OD_450_ was determined at 450 nm, and each sample was tested in duplicate. Corresponding non-specific binding was subtracted from the mean OD_450_ for each sample. The specificity of detection against lipid-free and unmodified apoA-1 has been previously determined by conventional saturation tests, Western blot, and LC-MS analyses (22). At an intermediate ratio of 0.6 OD_450_, the interassay coefficient of variation was 9% (n = 5), and the intra-assay CV was 5% (n = 5). For serum, the anti-apoA-1 IgG seropositivity cut-off was previously specified and validated, and was set at an OD_450_ ratio >0.64 for the 97.5^th^ percentile of anti-apoA-1 IgG of healthy blood donors (21, 22, 25-28, 38).

### IgG against Spike/apoA-1 and Spike/TLR2 mimic peptides assessment

The same protocol used for anti-apoA-1 IgG assessment was applied for the assessment of IgG against these two mimic peptides, *mutatis mutandis*, where they were used as coating antigen in our ELISA plates followed by our conventional protocol (21, 22, 25-28, 38).

### Cytokines and anti-pneumococcal IgG (P14 serotype) assessment

Interleukin 6 (IL-6), Tumor Necrosis Factor (TNF)-α, Monocyte Chemoattractant Protein (MCP)-1, interferon gamma (IFN)-γ and interferon α2a (IFN)-α2a were measured using the MesoScale Discovery (MSD) platform (Rockville, MD, USA) on the SQ120 instrument. Analyte concentrations were determined with Discovery Workbench^®^ software 4.0, using a 4 parameter logistic fit model. The lower limits of detection (LLOD) in pg/ml, the intra-run CVs and inter-run CVs for each cytokine were as follows: IL-6 (LLOD: 2.91 ; CV-intra-run: 1.23%; CV-inter-run: 9.8%), TNF-α (LLOD: 1.01 ; CV-intra-run: 3%; CV-inter-run: 8%), MCP-1 (LLOD: 1.43 ; CV-intra-run: 0.5%; CV-inter-run: 8%), IFN-γ (LLOD: 64.5 ; CV-intra-run: 0.3%; CV-inter-run: 6%) and IFN-α 2a (LLOD: 10.5 ; CV-intra-run: 4.9%; CV-inter-run: 0.25%).

As a negative control for anti-SARS-CoV-2 and anti-apoA-1-related serologies, IgG against pneumococcus belonging to the P14 serotype, the most prevalent serotype in Switzerland (52), was assessed for the derivation cohort only, using a SQ120 instrument (MSD platform) according to the manufacturers’ instructions. The seropositivity cut-off was 0.3 mg/l and the intra-run CV was 3.8% and inter-run CV was 12%.

### Cross reactivity and competition experiments

To assess the degree of cross reactivity between anti-SARS-CoV-2 and anti-apoA-1 IgGwith their respective antigens two kind of competition experiments were performed.

In the first one, anti-apoA-1 and anti-SARS-CoV-2 IgG positive sera were preincubated for two hours with different concentrations (7.5, 22, 66 and 200 ug/mL) of Spike and c-ter apoA-1 peptide respectively. We have choosen to compete anti-SARS-CoV-2 IgG signal with the c-terminal part of apoA-1 protein being the main anti-apoA-1 IgG epitope found in patients as previously described (29, 30) and because this apoA-1 region share linear homology with the c-ter sequence identified in Spike as previously reported. C-ter apoA-1 peptide in (>95% purity) was obtained by chemical synthesis (29), and Spike protein was kindly provided by the institute of technology EPFL (Lausanne, Switzerland). After the preincubation time, serum was added to a Spike or native and lipid-free apoA1 coated 96-well plates (Maxisorp plates, NuncTM, Roskilde, Denmark) according to an extensively validatedanti-apoA-1 IgG ELISA protocol, allowing the detection of antibodies against native and lipid-free apoA-1 (17, 21, 22, 25-27, 38).

The second competition experiment set was done using again our anti-apoA-1 IgG elisa protocol (21, 22, 25-28, 38), coating the plate with lipid-free apoA-1 or Spike protein and then increasing concentrations (5 to 40 µg/ml) of commercial polyclonal anti-Spike IgG (Thermo Fisher Scientific, Waltham, MA, USA, ref: PA5-81795) or polyclonal anti-apoA-1 IgG (Academy Biomedical Company, Houston, TX, USA, ref: 11A-G2) and their respective control antibodies, goat IgG (Meridian Life Science, Memphis, TN, USA, ref: A66200H) or rabbit IgG (Dako, Santa Clara, CA, USA, ref: X0903) were added to the plate for 30 min at 37 °C. Subsequently pooled sera from seven anti-apoA-1 and anti-Spike IgG positive patients were incubated onto the plate for one hour at 37°C. ApoA1 and Spike immunoreactivity was detected using an alkaline phosphatase-conjugated anti-human IgG as described above.

### Statistics

Results were reported as proportion, median, range and interquartile range (IQR), unless stated otherwise. Differences between groups were calculated using Fisher’s exact bilateral test (or Yates’ Chi-Squared test when appropriate) or the Mann-Whitney test for categorical and continuous variables, respectively. Spearman correlation was used to assess correlations between the variables. Receiving operating characteristics (ROC) analyses were used to determine the association with patients’ prognosis on the validation cohort according to the area under the curve (AUC). Cox regression analyses were used to determine the associations between pre-pandemic and post pandemic anti-apoA-1 IgG status in unadjusted mode, and after adjustment for age, gender and smoking. The same univariate analyses were used to determine whether pre-pandemic serology status could predict subsequent anti-SARS-CoV-2 serology status. Results were expressed as hazard ratios (HR) with 95% confidence intervals (95%CI). Due to the exploratory nature of this work and the pre-specified analysis planned, adjustments for multiple tests were not performed. All analyses were performed using Statistica software (version 13.5.0.17, TIBCO Software Inc., Palo Alto, CA, USA). Statistical significance was defined as *P* < 0.05. For *in vitro* studies, statistical analyses were performed using the Anova-test or *T*-test when two independent groups were analyzed using Prism 8 (GraphPad Software, San Diego, CA, USA). A 2-sided *P*-value < 0.05 was considered statistically significant.

## Data Availability

The data that support the findings of this study are available from the corresponding
author upon reasonable request.

## ACKNOWLEDGMENTS

The authors gratefully acknowledge Prof. Cosson from the Geneva Antibody Facility for generous gift of reagents, Isabelle Arm-Vernez from the virology laboratory of the laboratory medicine division for the assessment of routine SARS-CoV-2 IgG serology, as well as Julien Virzi, Diego Andrey and Patrick Cohen from the laboratory medicine division. We are grateful to Dr. Florence Pojer from the Protein Production and Structure Core Facility of the EPFL institute of technology of Lausanne who kindly provided purified Spike protein. Finally, we are indebted to Erik Boehm for his help in editing the manuscript and careful English revision.

## Author contributions

Conceptualization: NV, NW. Data curation : SP, GP, NS, SS, NW, SB, GP, CLT, JP, IG, LK. Formal analysis: NV, GP, SP, SS, GP. Funding acquisition: NV, SP, IG, LK, SS, JP. Investigation: SP, CJ, BM, LFS, SB, SY, BL, JPD, SB. Project administration: NV, SP, SY, LK, IG, NW. Resources: JP, NS, CLT, SY, LK, IE, SS, IG, OH, NW, JP, CSE, CAS, IE. Software: NS, CLT, GP, IG, SS. Supervision: NV, SP, NW. Validation: NV, SP, SY, LK. Visualization: SP, NW. Writing - original draft: NV, NW, SP. Writing - review & editing: All.

