## Supplemental figures and tables for "SARS-CoV2- infection as a trigger of humoral response against apolipoprotein A-1"

### Supplementary figures

### Supplementary fig. 1.

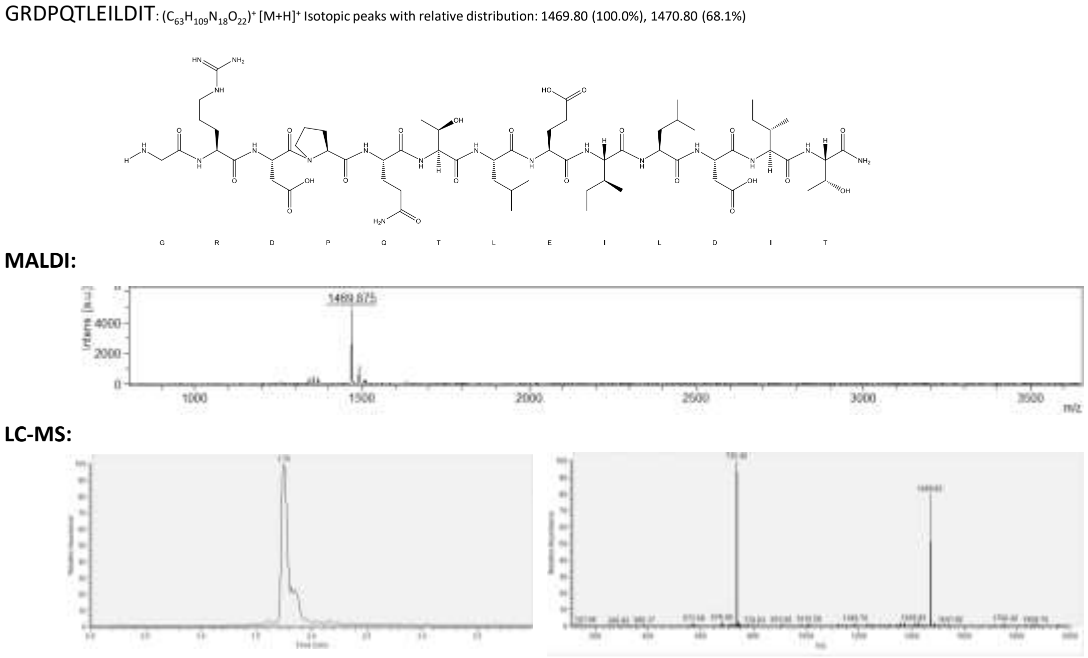


**Supplementary fig. 1. Spike-TLR2 mimic peptide (aa577-588 Spike protein SARS-CoV-2).**

GRDPQTLEILDIT amino acids sequence of the peptide Spike-TLR2 mimic. The peptide was purified by HPLC and the respective masses were verified by mass spectroscopy (MALDI).

**Supplementary fig. 2.**

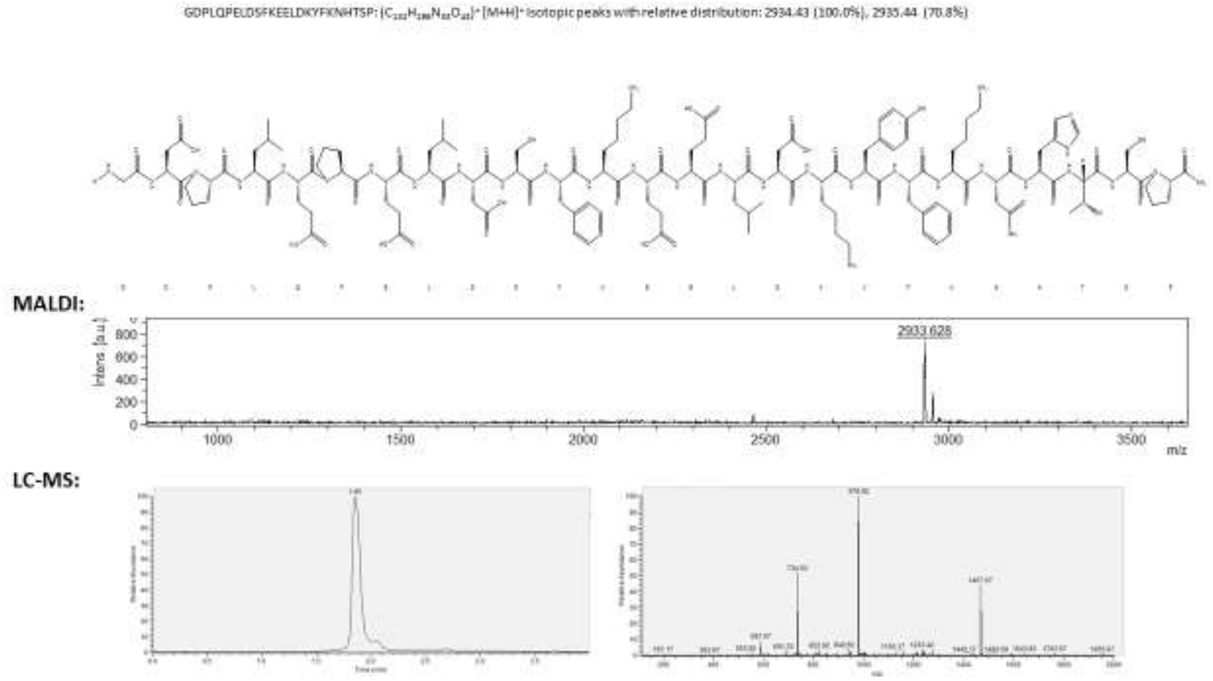


**Supplementary fig. 2. Spike-apoA-1 mimic peptide (1139-1162 Spike protein SARS-CoV-2):**

GDPLQPELDSFKEELDKYFKNHTSP amino acids sequence of the peptide Spike-apoA-1 mimic. The peptide was purified by HPLC and the respective masses were verified by mass spectroscopy (MALDI).

**Supplementary fig. 3.**

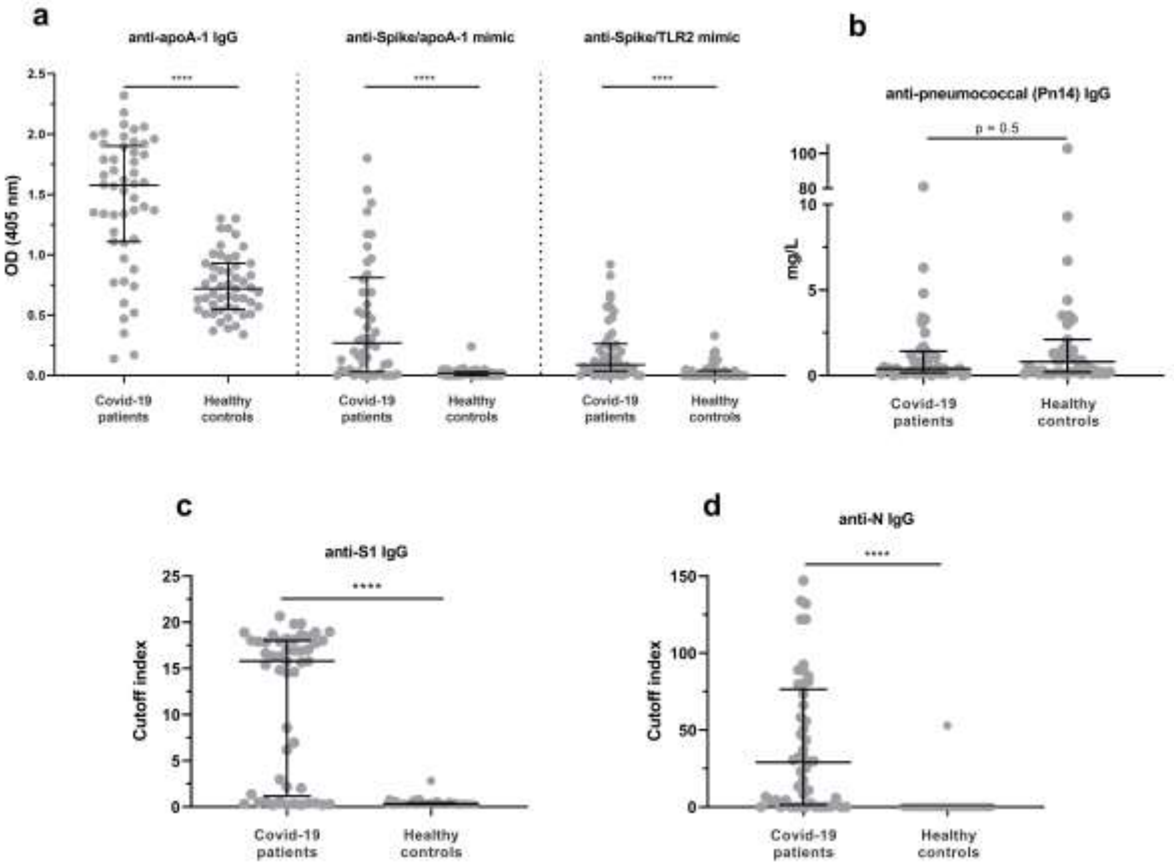


**Supplementary fig. 3. Levels of IgG against apoA-1, mimic peptides, SARS-CoV2 and p14 pneumococcus serotype in COVID-19 patients versus healthy donors (derivation cohort)**

**Panel a)**. Significant differences in antibody titers are observed between COVID-19 patients and healthy donors for anti-apoA-1 IgG (*****P* <0.0001), anti-Spike/apoA-1 mimic IgG (*****P* <0.0001) and anti-Spike/TLR2 mimic IgG (*****P* <0.0001) however **panel b)** shows that anti-pneumococcal (Pn14) antibody response doesn't change between the two groups (*P*=0.5). **Panel c)** shows a significant difference between COVID-19 patients and healthy donors for anti-S1 IgG (*****P* <0.0001) and panel d) presents the significant difference for anti-N IgG (*****P* <0.0001) between the two groups. Results are expressed as median with interquartile range and *P*-value calculated with Mann-Whitney test.

### Supplementary Tables

**Supplementary table 1. Spearman correlations between serologies and cytokines profile in the COVID-19 cases of the derivation cohort**

| | **Anti-S1 IgG** r ; *P*-value | **Anti-N total ab** r ; *P*-value | **Anti-apoA-1 IgG** r ; *P*-value | **Anti-Spike/apoA-1 mimic IgG** r ; *P*-value | **Anti-Spike/TLR2 mimic IgG** r ; *P*-value | **Anti-pneumococcal IgG** r ; *P*-value |
|---|---|---|---|---|---|---|
| ***Serologies*** | | | | | | |
| Anti-S1 IgG ; ratio | ND | 0.70 ;<0.0001 | 0.60 ;<0.0001 | 0.61 ;<0.0001 | 0.64 ;<0.0001 | -0.07 ; 0.50 |
| Anti-N total ab; ratio | 0.70 ; <0.0001 | ND | 0.54 ;<0.0001 | 0.62 ;<0.0001 | 0.40 ;<0.0001 | 0.07 ; 0.47 |
| Anti-apoA-1 IgG, $OD_{450}$ | 0.60 ; <0.0001 | 0.54 ;<0.0001 | ND | 0.60 ;<0.0001 | 0.54 ;<0.0001 | -0.03 ; 0.77 |
| Anti-Spike/apoA-1 IgG,$OD_{450}$ | 0.61 ; <0.0001 | 0.62 ;<0.0001 | 0.60 ;<0.0001 | ND | 0.50 ;<0.0001 | -0.03 ; 0.73 |
| Anti-Spike/TLR2 IgG,$OD_{450}$ | 0.64 ; <0.0001 | 0.40 ;<0.0001 | 0.54 ;<0.0001 | 0.50 ;<0.0001 | ND | 0.02 ; 0.84 |
| Anti-pneumococcal (Pn14) IgG, pg/ml | -0.07 ; 0.50 | 0.07 ; 0.47 | -0.03 ; 0.77 | -0.03 ; 0.73 | 0.02 ; 0.84 | ND |
| ***Cytokines*** | | | | | | |
| IFN-γ, pg/ml | 0.15 ; 0.14 | 0.12 ; 0.25 | -0.05 ; 0.63 | 0.22 ; 0.03 | 0.04 ; 0.70 | 0.01 ; 0.91 |
| IL-6, pg/ml | 0.41 ; <0.0001 | 0.45 ; <0.0001 | 0.36 ;<0.0001 | 0.35 ; 0.0003 | 0.29 ; 0.002 | -0.05 ; 0.59 |
| TNF-α, pg/ml | 0.38 ; 0.0001 | 0.40 ; <0.0001 | 0.26 ; 0.001 | 0.60 ;<0.0001 | 0.25 ; 0.01 | -0.07 ; 0.47 |
| MCP-1, pg/ml | 0.30 ; 0.003 | 0.14 ; 0.18 | -0.05 ; 0.73 | 0.35 ; 0.0004 | 0.26 ; 0.008 | 0.09 ; 0.36 |
| IFN-α2a, pg/ml | 0.43 ; <0.0001 | 0.37 ; 0.0001 | 0.26 ; 0.007 | 0.44 ;<0.0001 | 0.35 ; 0.0004 | -0.05 ; 0.62 |

ND: not determined; Anti-N total ab: anti-N antigen total antibodies

**Supplementary table 2. Baseline demographic and biological characteristics of ICU patients according to anti-S1 IgG status**

| Demographic and biological characteristics | Anti-S1 IgG seropositive patients (n=46) | Anti-S1 IgG seronegative patients (n=80) | *P*-value |
|---|---|---|---|
| Age, years | 62.5 (55-71;37-83) | 65.0 (57-73; 25-86) | 0.68 |
| Female gender; % (n) | 21.7 (10) | 22.5 (18) | 1.00 |
| BMI, kg/m2 | 27.7 (24.2-30.4;19.3-52.4) | 29.0 (25.7-32.3; 15.6-50.8) | 0.14 |
| Current smoking; % (n) | 13.0 (6) | 13.8 (11) | 1.00 |
| DPSO and ICU admission | 10.0 (8-13;3-27) | 8.0 (6-10;0-27) | 0.001 |
| ***Comorbidities*** | | | |
| Hypertension; % (n) | 45.6 (21) | 48.8 (39) | 0.85 |
| Dyslipidemia; % (n) | 28.2 (13) | 27.5 (22) | 1.00 |
| Diabetes; % (n) | 23.9 (11) | 28.8 (23) | 0.68 |
| Previous IC and or HF, % (n) | 21.7 (10) | 25.0 (20) | 0.83 |
| Previous stroke; % (n) | 4.3 (2) | 6.3 (5) | 1.00 |
| Known malignancy; % (n) | 4.3 (2) | 10.0 (8) | 0.32 |
| Chronic kidney disease; % (n) | 0 (0) | 11.3 (9) | 0.01 |
| ***Severity upon admission*** | | | |
| APACHE II score | 22 (12-26; 4-37) | 22 (15-29; 3-38) | 0.38 |
| SOFA score | 5 (4-7; 2-10) | 6 (4-7, 1-11) | 0.66 |
| SAPS II score | 54 (46-66; 18-81) | 52 (39.5-64; 6-82) | 0.52 |
| 28-days mortality,% (n) | 10.9 (5) | 20.0 (16) | 0.33 |
| Length of stay at ICU; days | 14.0 (8-18; 1-48) | 16.5 (11.5-22.5; 1-42) | 0.03 |
| Mechanical ventilation;% (n) | 91.3 (42) | 98.8 (79) | 0.06 |
| ***Cytokines and inflammation*** | | | |
| CRP, mg/l | 158.0 (111-224,24.4-402.8) | 150 (88.9-201; 23.1-311) | 0.22 |
| IFN-γ, pg/ml | 247.7 (104.8-691.5; 5.2-10180) | 450.9 (190.7-1155.1; 15.8-37747) | 0.02 |
| IL-6, pg/ml | 170.5 (86.4-511.1;6.7-7890) | 152.4 868.3-314; 13.9-2047) | 0.53 |
| TNF-α, pg/ml | 7.0 (3.9-12.6; 2.4-71.2) | 6.9 (4.4-17.5, 255-36484) | 0.71 |

| | | | |
|---|---|---|---|
| MCP-1, pg/ml | 5573 (2510-9212;776-30256) | 3813 82632-7535;255-36483) | 0.43 |
| IFN-α2a, pg/ml | 2.9 (1.2-13.6; 0.2-173.6) | 10.1 (3.9-31.4; 0.4-468.5) | 0.0005 |
| D-dimers; ng/ml | 1714.5 (1185-3129; 725-10001) | 1382 (858-2258; 220-9999) | 0.03 |
| ***Lipid profile*** | | | |
| Total cholesterol, mmol/l | 2.9 (2.5-3.4;0.2-1.0) | 2.6 (2.2-3.2; 1.1-4.7) | 0.25 |
| HDL cholesterol, mmol/l | 0.58 (0.44-0.68;0.17-1.00) | 0.64 (0.55-0.76; 0.14-1.95) | 0.03 |
| LDL cholesterol, mmol/l | 1.43 (1.02-1.79;0.38-3.72) | 1.21 (0.73-1.70; 0.00-3.1) | 0.12 |
| Triglycerides, mmol/l | 1.74 (1.36-2.52; 0.81-4.00) | 1.55 (1.11-2.17; 0.8-4.05) | 0.09 |
| ***Cardiac biomarkers*** | | | |
| Hs-cTnT, ng/L | 5.3 (1.6-15.9;1.1-27.0) | 19.2 (9.9-44.4-3.3-289) | 0.35 |
| NT-proBNP, pg/ml | 245 (131;898; 21.6-3657) | 310 (92.1-1220; 15.1-18772) | 0.95 |
| ***Serologies*** | | | |
| Anti-S1 IgG ; ratio | 5.3 (1.6-15.9;1.1-27.0) | 0.5 (0.4-0.7;0.3-1.1) | <0.0001 |
| Anti-N total ab; ratio | 5.95 (1.3-12.3; 0.1-36.3) | 0.12 (0.10.7;0.1-14.5) | <0.0001 |
| Anti-N total ab seropositivity;% (n) | 76.0 (35) | 22.5 (18) | <0.0001 |
| Anti-apoA-1 IgG seropositivity; % (n) | 41.3 (19) | 18.8 (15) | 0.01 |
| Anti-apoA-1 IgG, $OD_{450}$ | 0.58 (0.35-1.5; 0.00-2.60) | 0.37 (0.19-0.57;0.00-1.48) | <0.0001 |
| Anti-Spike/apoA-1 IgG, $OD_{450}$ | 0.08 (0.04-0.44;0.2.71) | 0.04 (0.02-0.07; 0.00-1.29) | <0.0001 |
| Anti-Spike/TLR2 IgG, $OD_{450}$ | 0.04 (0.02-0.09; 0-0.77) | 0.03 (0.01-0.07; 0-0.39) | 0.02 |
| ***Renal function*** | | | |
| Creatinine; µmol/l | 76 (66.5-105; 42-279) | 82.5 (66.5-105.5; 38-769) | 0.47 |

Anti-N total ab: anti-N antigen total antibodies

All continuous variables are expressed as median (interquartile range; and range).

**Supplementary table 3.** **Spearman correlations between serologies, cytokines, and lipid profile in the ICU patients**

| | **Anti-S1 IgG** r ; *P*-value | **Anti-N total ab** r ; *P*-value | **Anti-apoA-1 IgG** r ; *P*-value | **Anti-Spike/apoA-1 mimic IgG** r ; *P*-value | **Anti-Spike/TLR2 mimic IgG** r ; *P*-value |
|---|---|---|---|---|---|

| | | | | | |
|---|---|---|---|---|---|
| ***Serologies*** | | | | | |
| Anti-S1 IgG ; ratio | ND | 0.77 ; <0.0001 | 0.43 ; <0.0001 | 0.42 ; <0.0001 | 0.25 ; 0.005 |
| Anti-N total ab; ratio | 0.77 ; <0.0001 | ND | 0.44 ; <0.0001 | 0.53 ; <0.0001 | 0.24 ; 0.006 |
| Anti-apoA-1 IgG, $OD_{450}$ | 0.43 ; <0.0001 | 0.44 ; <0.0001 | ND | 0.51 ;<0.0001 | 0.34 ; <0.0001 |
| Anti-Spike/apoA-1 IgG, $OD_{450}$ | 0.42 ; <0.0001 | 0.53 ; <0.0001 | 0.51 ; <0.0001 | ND | 0.39 ; <0.0001 |
| Anti-Spike/TLR2 IgG, $OD_{450}$ | 0.25 ; 0.005 | 0.39 ; <0.0001 | 0.34 ; 0.0001 | 0.39 ; <0.0001 | ND |
| ***Cytokines*** | | | | | |
| CRP; mg/l | 0.18 ; 0.05 | 0.16 ; 0.08 | 0.20 ; 0.02 | 0.09 ; 0.32 | 0.09 ; 0.31 |
| IFN-γ, pg/ml | -0.25 ; 0.005 | -0.25 ; 0.005 | -0.19 ; 0.04 | -0.25 ; 0.001 | -0.13 ; 0.16 |
| IL-6, pg/ml | -0.02 ; 0.81 | -0.07 ; 0.47 | 0.11 ; 0.24 | 0.00 ; 0.98 | 0.00; 0.95 |
| TNF-α, pg/ml | -0.02 ; 0.83 | -0.09 ; 0.32 | 0.12; 0.19 | -0.09 ; 0.28 | 0.05; 0.58 |
| MCP-1, pg/ml | 0.04; 0.69 | 0.006 ; 0.94 | 0.01 ; 0.93 | 0.01 ; 0.93 | -0.06; 0.50 |
| IFN-α2a, pg/ml | -0.36; 0.0003 | -0.44 ; <0.0001 | -0.30 ; 0.0005 | -0.30 ; 0.0005 | -0.11 ; 0.22 |
| D-dimers; ng/ml | 0.29 ; 0.004 | 0.11 ; 0.26 | 0.05 ; 0.63 | 0.05 ; 0.63 | -0.04; 0.66 |
| ***Lipid profile*** | | | | | |
| Total cholesterol, mmol/l | 0.12; 0.21 | 0.04 ; 0.70 | -0.16; 0.06 | 0.07; 0.43 | 0.04; 0.66 |
| HDL cholesterol, mmol/l | -0.26; 0.004 | -0.27 ; 0.003 | -0.37; <0.0001 | -0.30; 0.001 | -0.25; 0.006 |
| LDL cholesterol, mmol/l | 0.12; 0.21 | 0.09 ; 0.33 | -0.16; 0.08 | 0.07; 0.45 | -0.04; 0.70 |
| Triglycerides, mmol/l | 0.24; 0.008 | 0.10 ; 0.29 | 0.20; 0.03 | 0.26; 0.004 | 0.25; 0.006 |

### Supplementary table 4. Evolution of seropositivity rates within one week in ICU patients

| Seropositivity rates | Day 1 | Day 3 | Day 7 |
|---|---|---|---|
| **Anti-apoA-1 IgG; (n)** | 17.6 %<br>(9/51) | 56.8 %<br>(29/51) | 82.4 %*<br>(42/51) |
| **Anti-S1 IgG; (n)** | 31.4 %<br>(16/51) | 66.7 %<br>(34/51) | 98.0 %*<br>(50/51) |
| **Anti-N ab; (n)** | 37.3 %<br>(19/51) | 78.4 %<br>(40/51) | 92.2 % *<br>(47/51) |

**P*-value per serology between Day 1 and Day 7: ≤0.0001 according to bilateral exact Fisher test.

**Supplementary table 5. Prognostic accuracy of serologies and other biomarkers' for 28-day mortality (21 deaths) within the ICU.**

| Predictors | AUC (95%CI); *P*-value |
|---|---|
| ***Serologies*** | |
| Anti-S1 IgG ; ratio | 0.62 (0.48-0.76); 0.05 |
| Anti-N total ab; ratio | 0.59 (0.45-0.75); 0.09 |
| Anti-apoA-1 IgG, OD | 0.53 (0.38-0.68); 0.37 |
| Anti-Spike/apoA-1 IgG, OD | 0.51 (0.37-0.65); 0.45 |
| Anti-Spike/TLR2 IgG, OD | 0.64 (0.51-0.77); 0.02 |
| ***Clinical scores*** | |
| APACHE II score | 0.67 (0.53-0.82); 0.01 |
| SOFA score | 0.58 (0.44-0.71); 0.14 |
| SAPS II score | 0.74 (0.62-0.86); <0.0001 |
| ***Cytokines and inflammation*** | |
| CRP, mg/l | 0.64 (0.50-0.799; 0.05 |
| IFN-γ, pg/ml | 0.59 (0.43-0.74); 0.14 |
| IL-6, pg/ml | 0.60 (0.47-0.73); 0.06 |
| TNF-α, pg/ml | 0.68 (0.54-0.81); 0.004 |
| MCP-1, pg/ml | 0.64 (0.52-0.77); 0.01 |
| IFN-α2a, pg/ml | 0.66 (0.53-0.79); 0.007 |
| D-dimers, ng/ml | 0.64 (0.50-0.78); 0.03 |
| ***Cardiac biomarkers*** | |
| Hs-cTnT, ng/L | 0.72 (0.60-0.85); 0.0002 |
| NT-proBNP, pg/ml | 0.72 (0.59-0.84); 0.0003 |

**Supplementary table 6. Characteristics of the general population participants recruited in 2016-2018 and subsequently included in the SEROCoV-POP during the COVID-19 pandemic**

| **Baseline characteristics (2016-2018)** | **General population (*n* = 663)** |
|---|---|
| Age in years, mean (SD) | 46.65 (13.11) |
| Female gender | 55.2 (366) |
| ***Comorbidities*** | |
| Diabetes, % (n) [a] | 5.0 (33) |
| Hypertension, % (n) [b] | 23.5 (156) |
| Cardiovascular diseases, % (n) [c] | 3.6 (24) |
| Current smoker, % (n) | 17.5 (116) |
| Obesity, % (n) [d] | 8.2 (54) |
| ***Biochemical parameters*** | |
| Anti-apoA-1 IgG seropositivity, % (n) | 25 (166) |
| Median Anti-apoA-1 IgG levels, OD (IQR) | 0.27 (0.20-0.38) |
| Total cholesterol, mmol/L, mean (SD) | 5.23 (1.03) |
| HDL cholesterol, mmol/L, mean (SD) | 1.67 (0.49) |
| LDL cholesterol, mmol/L, mean (SD)[e] | 3.03 (0.25) |
| Triglycerides, mmol/L, mean (SD) | 1.16 (0.64) |

SD: Standard deviation; IQR: interquartile range

[a] Diabetes was defined as glycaemia ≥7mmol/L or self-reported diabetes or diabetes medication.

[b] Hypertension was defined as mean systolic and/or diastolic BP ≥140/90 mmHg or self-reported hypertension or presence of anti-hypertensive medication.

[c] Cardiovascular diseases included myocardial infarction, angina pectoris and cerebral or leg vascular obstruction.

[d] Obesity/overweight was defined as body mass index ≥30 kg/m$^2$.

[e] LDL calculated according to Friedwald formula

**Supplementary table 7. Anti-apoA-1 IgG status in eleven anti-S1 IgG true false-positive patients (positive in S1 but negative in rIFA and anti-N serologies)**

| | Anti-S1 IgG, $OD_{450}$ | Anti-S1 IgG Interp. | Anti-N, ratio | Anti-N Interp. | rIFA Interp. | AAA-1 IgG $OD_{450}$ | AAA-1 IgG Interp. |
|---|---|---|---|---|---|---|---|
| Patient 1 | 10.14 | Pos | 0.06 | Neg | Neg | 0.49 | Neg |
| Patient 2 | 8.81 | Pos | 0.07 | Neg | Neg | 0.40 | Neg |
| Patient 3 | 6.84 | Pos | 0.07 | Neg | Neg | 0.24 | Neg |
| Patient 4 | 3.97 | Pos | 0.06 | Neg | Neg | 0.48 | Neg |
| Patient 5 | 7.76 | Pos | 0.08 | Neg | Neg | 0.35 | Neg |
| Patient 6 | 8.37 | Pos | 0.08 | Neg | Neg | 0.53 | Neg |
| Patient 7 | 8.97 | Pos | 0.07 | Neg | Neg | 0.18 | Neg |
| Patient 8 | 8.82 | Pos | 0.06 | Neg | Neg | 0.38 | Neg |
| Patient 9 | 5.82 | Pos | 0.06 | Neg | Neg | 0.30 | Neg |
| Patient 10 | 10.68 | Pos | 0.07 | Neg | Neg | 0.44 | Neg |
| Patient 11 | 10.02 | Pos | 0.06 | Neg | Neg | 0.52 | Neg |

AAA-1: Anti-apoA-1 IgG

rIFA: recombinant immunofluorescence
